# Complexities of cerebral small vessel disease, blood pressure, and dementia relationship: new insights from genetics

**DOI:** 10.1101/2023.08.08.23293761

**Authors:** Muralidharan Sargurupremraj, Aicha Soumare, Joshua C. Bis, Ida Surakka, Tuuli Jurgenson, Pierre Joly, Maria J. Knol, Ruiqi Wang, Qiong Yang, Claudia L. Satizabal, Alexander Gudjonsson, Aniket Mishra, Vincent Bouteloup, Chia-Ling Phuah, Cornelia M. van Duijn, Carlos Cruchaga, Carole Dufouil, Geneviève Chêne, Oscar Lopez, Bruce M. Psaty, Christophe Tzourio, Philippe Amouyel, Hieab H. Adams, Hélène Jacqmin-Gadda, Mohammad Arfan Ikram, Vilmundur Gudnason, Lili Milani, Bendik S. Winsvold, Kristian Hveem, Paul M. Matthews, WT Longstreth, Sudha Seshadri, Lenore J. Launer, Stéphanie Debette

## Abstract

**Importance:** There is increasing recognition that vascular disease, which can be treated, is a key contributor to dementia risk. However, the contribution of specific markers of vascular disease is unclear and, as a consequence, optimal prevention strategies remain unclear.

**Objective:** To disentangle the causal relation of several key vascular traits to dementia risk: (i) white matter hyperintensity (WMH) burden, a highly prevalent imaging marker of covert cerebral small vessel disease (cSVD); (ii) clinical stroke; and (iii) blood pressure (BP), the leading risk factor for cSVD and stroke, for which efficient therapies exist. To account for potential epidemiological biases inherent to late-onset conditions like dementia.

**Design, Setting, and Participants:** This study first explored the association of genetically determined WMH, BP levels and stroke risk with AD using summary-level data from large genome-wide association studies (GWASs) in a two-sample Mendelian randomization (MR) framework. Second, leveraging individual-level data from large longitudinal population-based cohorts and biobanks with prospective dementia surveillance, the association of weighted genetic risk scores (wGRSs) for WMH, BP, and stroke with incident all-cause-dementia was explored using Cox-proportional hazard and multi-state models. The data analysis was performed from July 26, 2020, through July 24, 2022.

**Exposures:** Genetically determined levels of WMH volume and BP (systolic, diastolic and pulse blood pressures) and genetic liability to stroke.

**Main outcomes and measures:** The summary-level MR analyses focused on the outcomes from GWAS of clinically diagnosed AD (n-cases=21,982) and GWAS additionally including self-reported parental history of dementia as a proxy for AD diagnosis (AD_meta_, n-cases=53,042). For the longitudinal analyses, individual-level data of 157,698 participants with 10,699 incident all-cause-dementia were studied, exploring AD, vascular or mixed dementia in secondary analyses.

**Results:** In the two-sample MR analyses, WMH showed strong evidence for a causal association with increased risk of AD_meta_ (OR, 1.16; 95%CI:1.05-1.28; P=.003) and AD (OR, 1.28; 95%CI:1.07-1.53; P=.008), after accounting for genetically determined pulse pressure for the latter. Genetically predicted BP traits showed evidence for a protective association with both clinically defined AD and AD_meta_, with evidence for confounding by shared genetic instruments. In longitudinal analyses the wGRSs for WMH, but not BP or stroke, showed suggestive association with incident all-cause-dementia (HR, 1.02; 95%CI:1.00-1.04; P=.06). BP and stroke wGRSs were strongly associated with mortality but there was no evidence for selective survival bias during follow-up. In secondary analyses, polygenic scores with more liberal instrument definition showed association of both WMH and stroke with all-cause-dementia, AD, and vascular or mixed dementia; associations of stroke, but not WMH, with dementia outcomes were markedly attenuated after adjusting for interim stroke.

**Conclusion:** These findings provide converging evidence that WMH is a leading vascular contributor to dementia risk, which may better capture the brain damage caused by BP (and other etiologies) than BP itself and should be targeted in priority for dementia prevention in the population.

**Key points:** *Question:* Do instrumental variable analyses leveraging genetic information provide evidence for a causal association of various vascular traits with Alzheimer’s disease (AD) and all-cause-dementia? How do these associations compare for white matter hyperintensity (WMH) burden, a highly prevalent marker of covert cerebral small vessel disease (cSVD), stroke, and blood pressure traits, the strongest known risk factor for cSVD and stroke?

*Findings:* Using Mendelian randomization (MR) leveraging large, published genome-wide association studies, this study showed a putative causal association of larger WMH burden with increased AD risk after accounting for pulse pressure effects, and some evidence for association of lower BP with AD risk with possible confounding by shared genetic instruments. Longitudinal analyses on individual-level data also supported association of genetically determined WMH with incident all-cause-dementia and AD, independently of interim stroke.

*Meaning:* This study using complementary genetic epidemiology approaches, identified increasing WMH burden to be associated with dementia and AD risk, suggesting the association as specific for cSVD and independent of BP and stroke.

## Introduction

As life expectancy rises worldwide, the prevalence of dementia is expected to reach 75 million by 2030.^1,2^ Devising strategies to prevent or delay its occurrence is therefore a major public health priority. As has been documented over decades, it is now widely recognized by the scientific community that a majority of dementia cases, including Alzheimer’s disease (AD) in the population are, in fact, due to a combination of vascular and neurodegenerative lesions.^3-6^ A large majority (80%) of patients with clinically diagnosed AD are found to have cerebrovascular lesions on post-mortem examinations.^7^ Epidemiological and clinical studies have shown that stroke patients have at least a doubling of their risk for incident dementia.^8,9^ At the population level, covert cerebral small vessel disease (cSVD), detectable on brain imaging in the absence of clinical stroke, is thought to be the main pathological substrate underlying the vascular contribution to cognitive decline and dementia,^10^ with nearly half of dementia cases exhibiting overlap of AD neuropathology with cSVD.^11^

White matter hyperintensity burden (WMH) is the most common magnetic resonance imaging (MRI) feature of covert cSVD. Evidence from observational studies has established strong associations of WMH with increased stroke and dementia risk, including AD,^12^ yet with no proof of causality. Recently, a putative causal relation between WMH and AD has been suggested in a preliminary Mendelian randomization (MR) analysis that used genetic instruments as proxies for WMH volume, thus leveraging the natural randomization of genetic variation at conception to mitigate risks of confounding and reverse causation as seen in observational studies.^13^ Intriguingly, however, while high blood pressure is by far the strongest risk factor for WMH and easily accessible to efficient therapy, several MR studies^14^ have reported inverse associations of genetically predicted blood pressure levels^15^ with AD. These associations were observed both in datasets using standard AD diagnostic criteria (IGAP)^16-18^ and in studies using self-reported parental history as a proxy for AD diagnosis (UK Biobank).^19^ “Selective survivorship”^20^ or age-dependent structural changes (arterial stiffness)^21^ and neurodegenerative lesions in blood pressure regulated regions resulting in reverse causation have been discussed as possible hypotheses.^22,23^ These inconsistencies have led to many discussions about the causal role of vascular damage in AD, but an understanding of the contributions of specific vascular pathology is still limited. Indeed, understanding such causal relationships is crucial to prioritize interventions and target populations to prevent cognitive decline and dementia.

Based on the sharing of a large proportion of genetic risk variants between WMH and blood pressure (BP) traits,^24^ we aim to address from a genetic epidemiologic perspective: 1) what are the putative causal associations of genetically defined different vascular pathologies to AD and all-cause-dementia; 2) what are the potential biases that may contribute to the MR associations.

## Methods

The study design is summarized in **Figure 1**.

**Figure 1:**
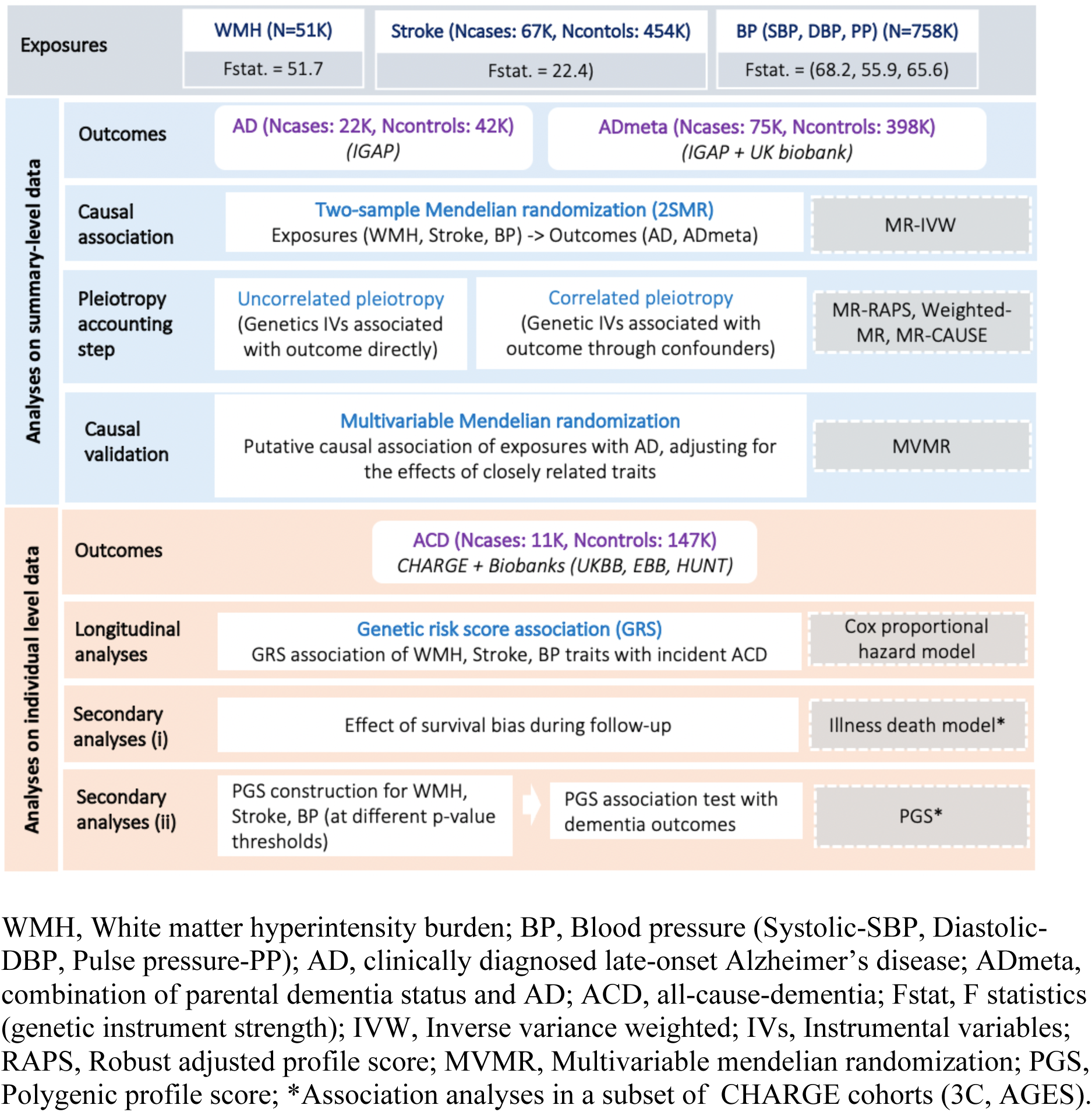
Study workflow.

### Analyses on summary level data

A comprehensive suite of MR-based analyses leveraging summary statistics of large GWAS was used to establish causality of associations between vascular traits (WMH, BP, stroke) and dementia and to test for alternative interpretations of causality. A *P* value < .017 correcting for 3 independent traits was considered significant.

#### Selection of genetic instruments for the vascular trait exposures in MR analyses

Independent genetic variants (single-nucleotide polymorphisms [SNPs]) that are genome-wide significant (P < 5×10^-8^) in large published GWAS and satisfying the instrumental variable definition^25,26^ were considered as genetic instruments for each of the exposures. These were derived from GWAS of WMH on 50,970 participants, of stroke on 67,162/454,450 cases and controls, and of systolic blood pressure [SBP], diastolic blood pressure [DBP], and pulse pressure [PP] on up to 757,601 participants.^15,24,27^ (**eMethods**). The Cragg-Donald F statistic was used to evaluate instrument strength (**eMethods**).

#### Dementia phenotypes used as outcomes in MR analyses

For the MR analyses, we used the GWAS summary statistics of clinically diagnosed late-onset AD (N=21,982/41,944 cases/controls)^28^ and of the combination of clinical AD cases (n-cases=21,982) and proxy AD cases (n-cases=53,402) defined by parental history of dementia in the UK biobank (this combination being hereafter referred to as AD_meta_, N=75,024/397844 cases/controls).^29^ AD_meta_ is sometimes considered closer to a definition of all-cause-dementia than pure AD.^30^ In all instances AD diagnosis was based on clinical criteria and family history only, and not on neuropathology criteria (**eMethods**).

#### Suite of MR analyses

As a starting point the putative causal effect of a given risk factor (WMH, stroke, SBP, DBP, PP) on AD was estimated using a suite of instrumental variable analyses based on two-sample Mendelian randomization (2SMR). We first accounted for potential pleiotropic effects of genetic instruments (SNPs) directly on the outcome and that is uncorrelated with the exposure (uncorrelated pleiotropy)^31^ using MR Egger, MR-RAPs, weighted median and mode based methods (**eMethods**). Second, we used MR-CAUSE to account for possible correlated pleiotropy, whereby genetic instruments are correlated with both exposure and outcome through the unmeasured confounders (**eFigures 1-2** and **eMethods**).^32^ MR-CAUSE is a Bayesian approach that differentiates the causal model (*γ*) from the correlated-sharing (q) model based on the extent of the contribution of genetic instruments to the predicted effect. It provides a prediction accuracy (expected log pointwise posterior density, ELPD) for both models and their corresponding difference (ΔELPD=ELPD_*γ*_-ELPD_q_), which, if greater than zero, indicates that the causal model (*γ*) is a better fit than the correlated-sharing model (q) (**eMethods**). Third, for exposures for which MR-CAUSE suggested the causal model to be a better fit (ΔELPD > 0) but with significant residual effects in the sharing model (q), we performed multivariable MR conditioning for effects from potential confounders to confirm the putative causal relation using MVMR.^33^ Finally, for exposures with significant MVMR association (*P* value < .013, for 4 independent traits) the following sensitivity analyses were conducted: i) using Qhet-MVMR, confounding due to potentially weak instruments was accounted for, and the effect direction was confirmed,^34^ and ii) causal direction among the multiple exposures was further determined using two-sample bidirectional MR (**eMethods**).

### Analyses on individual level data from longitudinal cohort studies

We additionally conducted analyses on individual level data from prospective cohort studies to study the relation of genetically determined WMH burden, BP, and stroke with incident dementia outcomes in a longitudinal setting, while exploring the impact of potential selective survival bias.^35^

#### Association analyses of weighted genetic risk scores for vascular traits with incident dementia

Analyses are based on genotype and phenotype information from 13 longitudinal cohorts (participating in the Cohorts for Heart and Aging Research in Genomic Epidemiology [CHARGE],^36^ and large biobanks (Trøndelag health study: HUNT, Estonian biobank: EstBB, and UK biobank: UKB). Nearly all cohorts were population-based, except MEMENTO (memory clinic patients with cognitive complaints and no dementia) and Knight ADRC. Together these cohorts included 157,698 unrelated participants of European origin, of whom 10,699 developed all-cause-dementia (which is more inclusive than AD_meta_). Dementia diagnosis in these studies is based on standard criteria described in the **eMethods** along with the study design and participant selection. The mean age at dementia diagnosis across the cohorts is 71.4 years with a median duration of follow-up ranging between 3-25 years. Cox proportional hazard regression models were used to examine the association of genetic risk scores for the different exposures (WMH, stroke, BP traits) studied in the 2SMR analyses with incident all-cause-dementia. For each trait, we constructed an individual weighted genetic risk score (wGRS) using as weights the number of ‘effect’ alleles times the effect estimates (betas) of the respective genetic instruments constructed for a given exposure in the 2SMR analysis.^37^ The wGRSs were standardized to have mean of 0 and a variance of 1, so that each unit change in the wGRS corresponds to one standard deviation (SD). Analyses were restricted to participants with at least one follow-up visit and no dementia at baseline. The Cox model with delayed entry included birth as time origin and age as the time scale and controlled for sex, education level, principal components of population stratification, and study-specific criteria (**eTable 3**). Data were censored at the age of dementia diagnosis for cases or age at the last follow-up for controls. Individual cohort-specific estimates were combined using a fixed-effects inverse variance weighted meta-analysis, implemented as an R (meta) package. Additional sensitivity analyses excluding individuals with a history of stroke at inclusion and adjusting for interim incident stroke were conducted (except in Knight ADRC where this information was not available).

#### Secondary analyses accounting for interval-censoring, competing risk of death, and dementia subtypes

These in-depth analyses were conducted in two large CHARGE cohorts (3C [Three city], AGES [Age, Gene/Environment Susceptibility - Reykjavik study]) with nearly identical characteristics (sample sizes, age distributions, and dementia ascertainment) (**eMethods**). We first explored whether survival bias during follow-up might affect our results using illness death models (IDMs)^38^ accounting for interval-censoring of time-to-onset of dementia and competing risk of death. Second, we examined whether our vascular exposures of interest exhibit an association with different subtypes of incident dementia (all-cause-dementia, AD and vascular and/or mixed dementia, **Supplementary Methods**) at more liberal instrument selection thresholds using polygenic profile scores (PGSs), created by binning observations by p-value of the exposures in the original GWASs. A *P* value < .017 correcting for 3 independent traits was considered significant.

## Results

### Exploring causal relations of WMH, BP, and stroke with AD risk using GWAS summary-level data

The genetic instruments derived from GWAS of WMH, BP, and stroke strongly predicted the exposures, with a Cragg-Donald F statistic ranging between 22 and 65 (**eTables 1-2**). Using the inverse variance weighting method we found significant associations (*P* value < .017) of genetically determined larger WMH burden and lower BP (DBP, SBP, PP) levels with AD_meta_ (clinically diagnosed AD or parental dementia) risk (**Figure 2, eTable 4a**) and of lower DBP with clinically diagnosed AD risk (**eFigure 3, eTable 5a**). The complementary MR tools MR-RAPS, weighted-median and mode enabled us to robustly rule out uncorrelated pleiotropic effects (**eTable 4b, 5b**). The Bayesian MR-CAUSE method that accounts for correlated pleiotropy (**Methods**) supported a causal relation of WMH with both AD outcomes (clinical AD and AD_meta_), with a posterior distribution of the causal model distinctively different from the sharing model (ΔELPD = 0.91 for AD; 0.50 for AD_meta_) (**eTable 6, eFigures 4-5**). On the contrary, stroke and BP traits suggested a better fit of the sharing model with potential unmeasured confounders (ΔELPD < 0) than a causal model (**eTable 6, eFigures 4-5**). Notably, for WMH with AD and not for other exposure-outcome combinations, we observed the presence of a significant proportion of genetic instruments being shared with unmeasured confounders (q *P* value: 8.37×10^-9^, **eFigure 5**) along with a better fit of the causal model (ΔELPD > 0) (**eTable 6**). We, therefore, sought to confirm any putative causal association of WMH with AD in a multivariable analysis setting, adjusting for the effects of closely related traits using MVMR. Greater genetically predicted WMH burden was associated with a 27.8% increase in the probability of AD risk (OR: 1.28, CI:1.07-1.53, *P*=.008, per unit increase in WMH risk alleles) after accounting for PP effects (**Figure 3**, **eTable 7**). This represents a 16.5% increase in disease risk compared to the univariable estimates (OR:1.11, CI:0.95-1.31*, P*=.19), with a consistent direction of effect. The effect of other exposures (stroke, BP traits) on AD remained non-significant using MVMR after adjusting for the closely related traits (**eTable 7**). Finally, a bidirectional MR analysis between the exposures showing significant MVMR results (WMH, PP) suggested a causal path of higher genetically predicted levels of PP with larger WMH burden (**eTable 8**).

**Figure 2:**
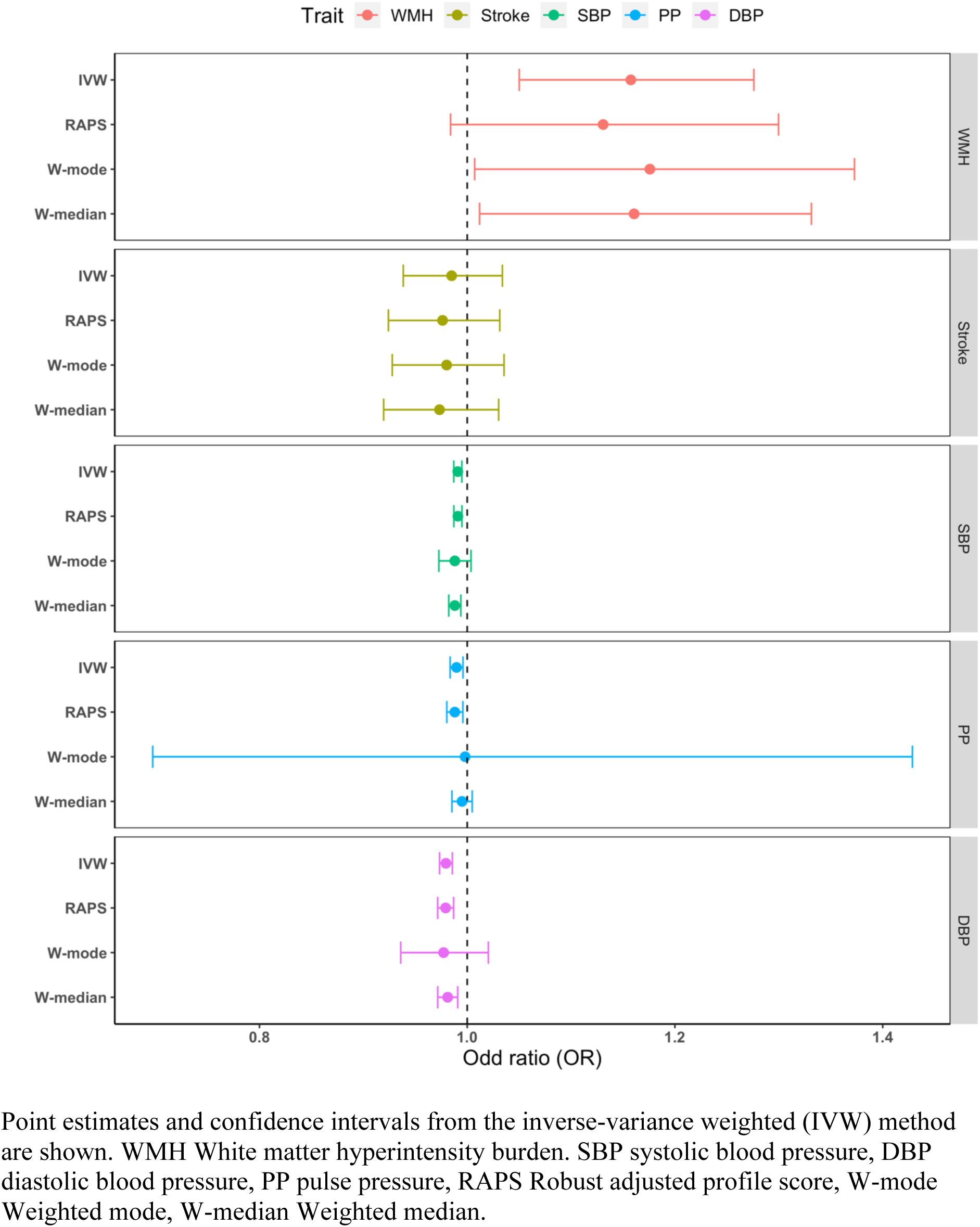
Mendelian randomization results of vascular risk factors with ADmeta.

**Figure 3:**
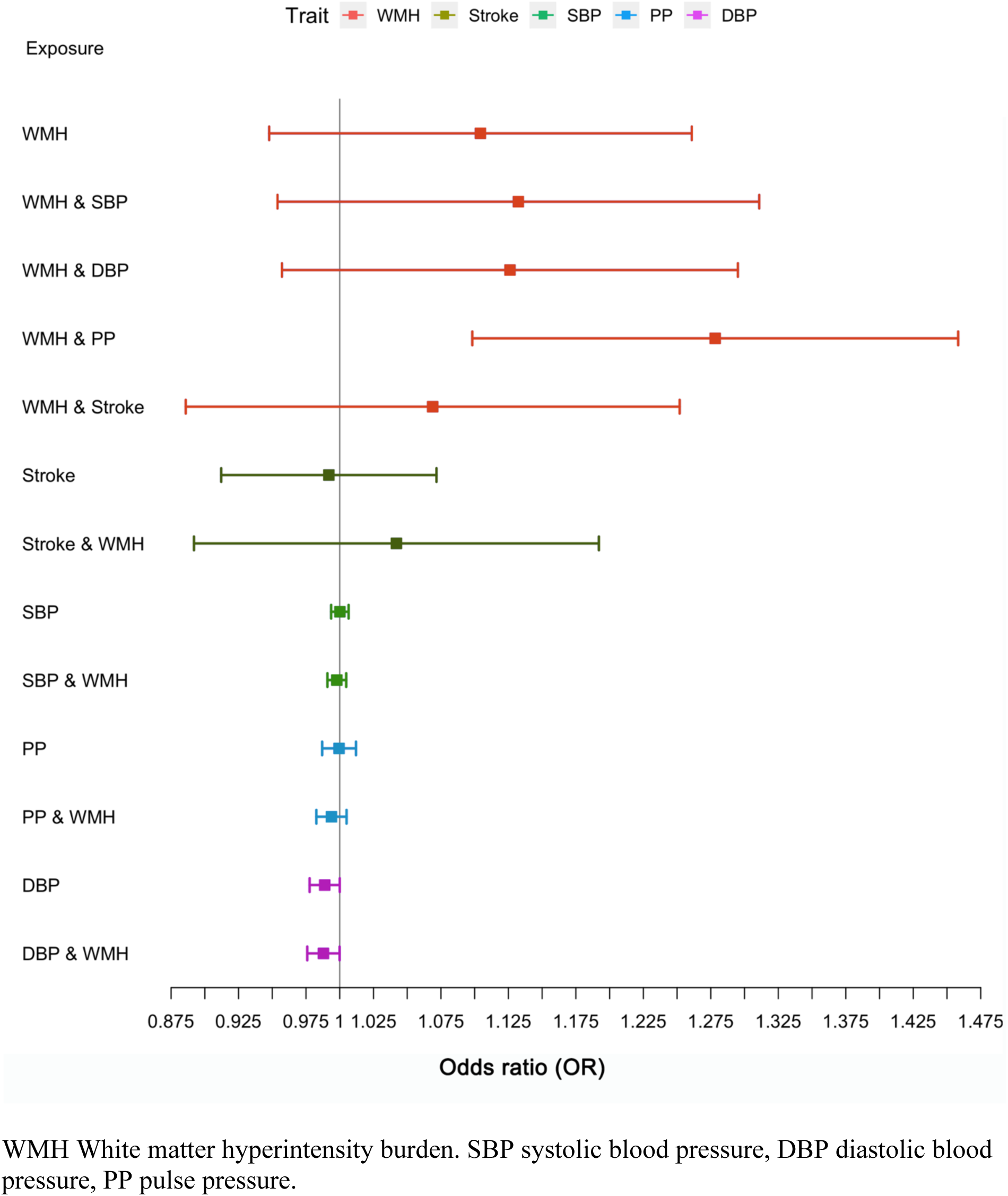
Multivariable Mendelian randomization (MVMR) along with the univariable MR for AD as the outcome.

### Association of genetic risk scores for WMH, BP, and stroke with incident dementia using individual-level data

In a meta-analysis of thirteen longitudinal cohort studies, we observed a borderline significant association of increasing genetically predicted WMH burden with increased risk of incident all-cause-dementia (HR:1.02, CI:1.00-1.04*, P*=.06, per SD increase in WMH wGRS) (**Figure 4, eTable 9**). After adjusting for education and interim stroke, this association remained substantially unchanged (**Figure 4**). There was no significant heterogeneity across cohorts (I^2^: 7%, p=0.38; **eFigure 6**). Genetically determined BP traits and genetic liability to stroke failed to show significant associations with incident all-cause-dementia, with negative point estimates for stroke and SBP. Notably, all exposures showed at least a nominally significant association with increased risk for mortality, the most significant association being observed for SBP (**eTable 9-10**).

**Figure 4:**
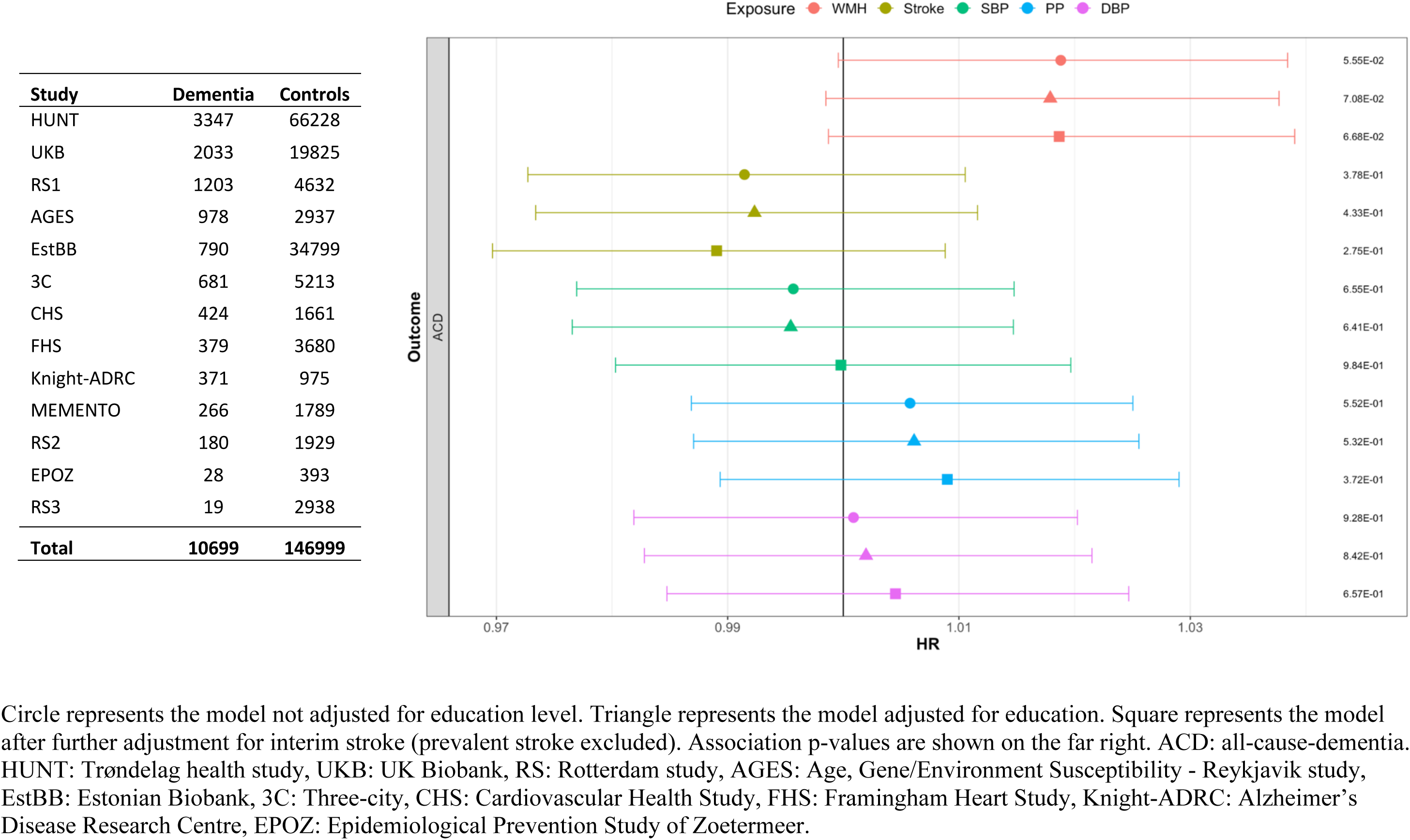
Forest plot showing the meta-analysis results of risk factor wGRSs (per standard deviation increase) with incident ACD.

In secondary analyses, using illness-death models (IDM) in two older population-based cohorts (3C, AGES, **Methods**), we ruled out potential biases related to competing risk of death during follow-up in the context of interval censoring. Indeed, per SD increase in genetically predicted WMH and BP levels, and genetic liability to stroke risk were not associated with incident all-cause-dementia in the IDM model, and the effect estimates were generally similar to those from the Cox model (**eTable 11**) in both cohorts.

When examining the PGSs, where associations are binned according to significance of genetic instruments with less stringent instrument-significance thresholds (**eMethods**), we found that PGSs for WMH and stroke were significantly associated (P < .017) with increased all-cause-dementia risk in both cohorts (**eTable 12-13**, **eFigures 7-8**). In sensitivity analyses excluding prevalent stroke and adjusting for interim stroke, the WMH PGSs associations with dementia outcomes remained unchanged, while the stroke PGSs associations with dementia risk were markedly attenuated in both cohorts (**eTable 13, eFigure 8**). Meta-analyses of effect estimates from individual PGSs bins in 3C and AGES showed a significant association of WMH and stroke PGSs with increased risk of all-cause-dementia, AD, and (for stroke) vascular and/or mixed dementia (**eTable 14**). Finally, most of the PGSs for BP traits failed to show significant associations with dementia risk; the significant protective association observed between SBP/DBP PGSs and AD in the AGES cohort was attenuated in sensitivity analyses excluding prevalent stroke and adjusting for interim stroke (**eTable 15**).

## Discussion

In this study capitalizing on a comprehensive MR workflow, large GWASs and numerous longitudinal cohort studies and biobanks, we report a putative causal association of genetically predicted WMH burden with increased risk of both clinically diagnosed AD^28^ and AD_meta_ (AD and parental history of dementia).^29^ The former association was strengthened after accounting for PP using multivariable MR. In contrast we observed protective effects of BP traits with AD_meta_ (all BP traits) and AD (DBP) risk using two-sample MR,^16-19^ and provide evidence that these may at least partly be driven by the sharing of BP genetic instruments with unmeasured confounders. Genetic liability to stroke was not associated with AD or AD_meta_. Next, using 13 large longitudinal cohorts and biobanks gathering 157,698 participants with prospective dementia surveillance (N=10,699 incident dementia cases), we report a similar, borderline significant association trend of WMH with increased risk of incident all-cause-dementia, while genetic instruments for BP traits and stroke showed no association. In a subset of population-based cohorts, polygenic scores (encompassing risk variants at less stringent significance thresholds than MR instruments) for WMH burden and stroke were associated with increased risk of incident all-cause-dementia, AD, and vascular and/or mixed dementia. While the association between stroke PGSs and all-cause-dementia attenuated markedly after adjusting for interim stroke, the WMH PGSs association with all-cause-dementia remained significant.

Overall, out of all the vascular phenotypes considered (WMH, BP traits, stroke), genetic instruments for WMH appear to show the most robust associations with dementia risk, including AD_meta_, AD, and all-cause-dementia. These MR findings suggest causal mechanisms and highlighting WMH as an important causal pathway to target for the prevention of dementia (**Figure 5**). Our results support association of extensive WMH burden with dementia and AD risks described in observational studies,^39-43^ providing additional strong evidence for a possible causal relation. This reinforces our earlier preliminary observations of a putative causal association of WMH with AD_meta_,^24^ and expands it to a larger AD_meta_ GWAS resource,^29^ and, importantly also to a smaller and more conservative definition of only clinically diagnosed AD.^28^ The fact that the latter association was more prominent after accounting for PP effects in a multivariable model, with a 16.5% increase in the disease risk compared to the univariable estimates, is intriguing. PP is a marker of the pulsatile component of BP and is correlated with measures of arterial stiffness,^44,45^ which was suggested to promote both white matter pathology and amyloid deposition.^46,47^ Interestingly, recent evidence for a causal relation of arterial stiffness with larger WMH burden was found to be reinforced after accounting for PP,^48^ in line with the present findings. Underlying mechanisms are speculative, but could potentially involve oxidative stress, implicated in BP-associated vascular damage^49^ and also an early and prominent feature of aging and neurodegeneration.^50^ Elevated PP may dysregulate brain endothelial cells and increase cellular production of oxidative and inflammatory molecules, which may in turn increased amyloid-β secretion by cerebral endothelial cells and induce blood-brain barrier breakdown.^51-53^ Interestingly, although WMH is known to be associated with increased risk of stroke,^12,13^ and stroke with a substantial increase in dementia incidence,^54-56^ associations of WMH PGSs with risk of dementia were unchanged after accounting for baseline and interim stroke, suggesting an independent effect of WMH on dementia risk.

**Figure 5:**
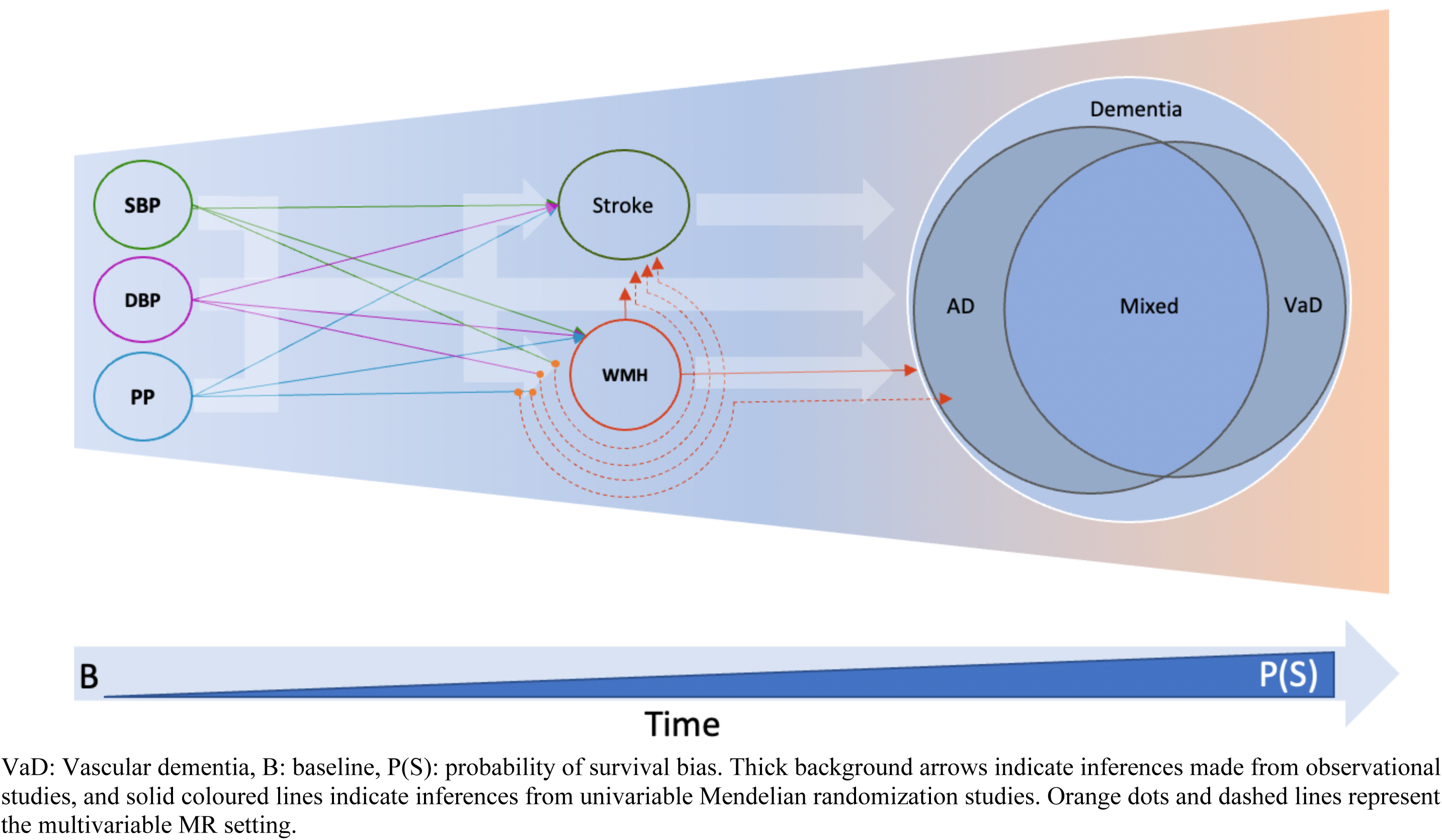
Central role of WMH with dementia outcomes.

High blood pressure is the strongest known risk factor for WMH and other MRI-markers of cSVD, with MR studies suggesting a causal relation of increasing genetically determined BP with WMH volume, even in persons without clinically defined hypertension.^24^ Moreover, evidence from randomized controlled trials shows that WMH volume progression is slowed down by BP-lowering treatments in hypertensive individuals,^57-61^ especially by intensive vs. standard BP-lowering.^61^ Given the aforementioned associations of genetically determined WMH with dementia and AD it therefore appears “counterintuitive” to observe an inverse association of genetically determined high BP with lower risk of AD_meta_ and AD in our two-sample MR analysis. It is however in line with earlier MR studies deriving instruments from earlier, smaller BP GWAS or using genetic proxies for the effect of BP-lowering drugs.^16-19,62^ Our sensitivity analyses suggest that this unexpected directionality of association might at least partly be explained by pleiotropic effects from unmeasured confounders (MR-CAUSE), prompting caution with applying methods designed only to remove or downweigh pleiotropic variants (e.g., weighted median based MR methods).

Moreover, while we saw suggestive evidence for selective survival during follow-up, given the late age of onset of dementia (85 years on average^63^), the strong association of high BP and stroke with premature death in our longitudinal data and extensive evidence from other studies,^64-67^ it is possible that the apparent protective effect of high BP on dementia risk could potentially reflect selective survival bias before entry into the study. Indeed, if the selection is a function of the exposure or the outcome, it could result in collider (index event) bias,^68^ leading to an association in the absence of a causal effect.^69-71^ While we did not see any evidence for selective survival during follow-up using the illness-death model in our longitudinal cohorts, we cannot formally rule out selective survival before study entry. Although non-significant, the association of genetically determined PP and DBP with incident all-cause-dementia had point estimates above 1 in the longitudinal cohort studies, which are likely less exposed to selective survival than the AD case-control GWAS used for the 2SMR analyses, although they also tend to overrepresent healthy individuals at inclusion (as for any voluntary participation).^72-74^ Beyond these possible biases our results highlight the complexity of the relation between BP and dementia risk, with weak epidemiological evidence varying by age.^75-77^ Recent meta-analyses of clinical trials show evidence that antihypertensive medication reduces risk of secondary outcomes combining dementia with cognitive impairment,^78,79^ but not dementia alone. In a meta-analysis of 6 population-based cohorts, antihypertensive medication was associated with a reduced risk of incident dementia and AD in individuals with clinically defined high BP at baseline.^78^ However in another recent meta-analysis of 7 population-based cohorts, dementia risk appeared lower for older individuals with higher SBP levels, with more distinctly U-shaped associations for participants older than 75 years, which were not explained by SBP-associated changes in mortality risk.^80^

In contrast with BP measurements, which show high intra-individual variability,^81^ WMH volume is also a more stable marker, reflecting white matter damage secondary to changes in structure or function of cerebral small vessels. Assuming that WMH at least partly mediates the association of BP with dementia in the population, WMH may better capture the brain damage caused by BP than BP itself. WMH and covert cSVD more broadly likely also reflects the impact of other parameters on white matter integrity, including other risk factors for cSVD such as cerebral amyloid angiopathy, or factors influencing the resilience of the brain white matter to vascular insults. Given the high prevalence of WMH in the general population in the absence of clinical stroke,^75-77^ our results highlight WMH as a major causal pathway to consider for the prevention of dementia (**Figure 5**).

## Strengths and Limitations

Strengths of our study include the diversity and complementarity of the analytical approaches used, ranging from various state-of-the-art MR analyses, based on powerful published GWAS, to longitudinal analyses in large population-based cohort studies with prospective dementia ascertainment. We also acknowledge limitations. First, the GWASs used for the MR analyses have likely predominantly recruited patients with clinically diagnosed AD, thus leading to an underrepresentation of patients with mixed dementia, who likely represent the majority of AD cases in the population. Second, in our longitudinal analyses, the number of incident dementia cases remained modest, with some differences in the methods for dementia ascertainment, thus limiting power to detect associations. Third, we considered only one MRI-marker for cSVD, WMH, for which there is most evidence for an association with dementia risk and the strongest genetic instruments. With increasing availability of larger GWAS for other MRI-cSVD markers (cerebral microbleeds, lacunes, perivascular spaces) future studies concomitantly assessing the impact of various genetically determined MRI-cSVD markers are warranted.^82-85^ Finally, validation of our findings in populations of non-European ancestry, as larger datasets become available, will be crucial.

## Conclusions

In summary, our findings provide converging evidence that WMH is a leading vascular contributor to dementia risk and should be targeted in priority for the prevention of cognitive decline and dementia in the population. They also support WMH as a relevant surrogate marker for clinical trials testing interventions to prevent dementia by better controlling vascular risk.^61,86^ Our data also prompt caution when interpreting MR studies where the outcome of interest is a late-onset disease such as dementia, especially if the instrument for the exposure of interest shows a strong association with survival. They highlight the importance of combining several complementary epidemiological approaches to compensate for respective limitations and of combining different studies, cohorts and biobanks rather than drawing definitive conclusions on single datasets to minimize the impact of study-specific shortcomings and biases.

## Data Availability

Analyses in the manuscript uses publicly available genetic association summary statistics. The longitudinal information from the individual cohorts is available for additional analysis through contacting and collaborating with the individual cohorts as detailed in the supplementary methods.

