## Supplementary Methods for "Complexities of cerebral small vessel disease, blood pressure, and dementia relationship: new insights from genetics"

**SUPPLEMENTARY ONLINE CONTENT**

**SUPPLEMENT 1**

**eMethods.**
Individual study level: study population and dementia diagnosis (pages 2-8)
Genetic instruments selection (page 8)
Mendelian randomization analyses (pages 8-10)
Longitudinal analyses (pages 10-11)

**eAppendix 1. Acknowledgements** (page 12)
**eAppendix 2. Conflict of Interest Disclosures** (page 12)

**eFigure 1. Analytical pipeline for the MR approaches.** (page 13)
**eFigure 2. Mendelian randomization model with different types of pleiotropy; and the included exposures and outcomes.** (page 13)
**eFigure 3. Mendelian randomization results of vascular risk factors with AD.** (page 14)
**eFigure 4. Comparison of the sharing and the causal model using CAUSE for ADmeta as the outcome.** (page 15)
**eFigure 5. Comparison of the sharing and the causal model using CAUSE for AD as the outcome.** (page 16)
**eFigure 6. Forest plot showing association of WMH-wGRS with incident all-cause dementia (study-wise).** (page 17)
**eFigure 7. WMH polygenic profile score (PGS) association with dementia outcomes in 3C (blue) and AGES (black).** (page 18)
**eFigure 8. Stroke polygenic profile score (PGS) association with dementia outcomes in 3C (blue) and AGES (black).** (page 19)

**eReferences** (pages 20-23)

### eMethods

#### Individual study level: study population and dementia diagnosis

Overall, dementia diagnosis in CHARGE cohorts and other longitudinal cohorts participating in the individual level data analyses were based on the Diagnostic and Statistical Manual of Mental Disorders using either DSM-III-R or DSM-IV. For AD, the National Institute of Neurologic and Communicative Disorders and Stroke–Alzheimer's Disease and Related Disorders Association (NINCDS-ADRDA) criteria were used, while vascular dementia was defined according to the National Institute of Neurological Disorders and Stroke (NINDS) and the Association Internationale pour la Recherche et l'Enseignement en Neurosciences (NINDS-AIREN) and State of California Alzheimer's Disease Diagnostic and Treatment Centers (ADDTC) criteria. Mixed dementia is defined as AD with vascular contributions.[26,27] For the biobanks dementia and AD cases were ascertained by the International Classification of Diseases (ICD9 and ICD10) coding system. Details on dementia ascertainment are provided in the Supplementary Methods.

**Three-city:** The protocol of the Three-City (3C) Study was approved by the Consultative Committee for the Protection of Persons Participating in Biomedical Research at Kremlin-Bicêtre University Hospital, Paris, France, and the Committee for the Protection of Persons, Sud-Mediterranée III, Nîmes, France. All participants provided written informed consent. The 3C Study is a prospective cohort initiated in 1999-2000 among 9294 noninstitutionalized community dwellers aged at least 65 years selected from the electoral rolls of 3 French cities (Bordeaux [n = 2104], Dijon [n = 4931], and Montpellier [n = 2259]).[1] Participants have been followed every 2 to 3 years during face-to-face interviews conducted at home and at the study center, with repeated cognitive evaluations and ascertainment of incident dementia cases until 2012 in Dijon and until 2016 in Bordeaux and Montpellier (date of final follow-up, July, 2016). Participants in the overall cohort were excluded if they had prevalent dementia at baseline, or if they were not followed up for dementia. Dementia status was evaluated prospectively by an expert panel. In Bordeaux and Montpellier, all participants were examined by a neurologist. In Dijon, due to the large number of participants, a two-step procedure was used:[1] (i) a careful neuropsychological evaluation carried out by a trained psychologist and (ii) an examination by a neurologist for those who screened positive at step 1 based on MMSE and Isaacs' Set Test, a measure of verbal fluency and response rapidity that consists of generating words belonging to given semantic categories (e.g., animal names) in 15 s.[1] Isaacs' Set Test has been reported to show the earliest decline in the decade preceding dementia diagnosis.[2,3] Cutoff scores were defined according to educational level as previously described.[4] Finally, in all centers, the diagnosing examination and subtype classification of all suspected prevalent and incident dementia cases were performed by an independent committee of neurologists following DSM-IV criteria. The final diagnosis of dementia and subtype was made based on all available information, including data on cognitive functioning and daily activities, severity of cognitive disorders and, where possible, hospitalization records, CT scans (which were most often used at the beginning of the follow-up period) and magnetic resonance images,[1] and functional assessment, which included assessment of disabilities using the Katz Index of Activities of Daily Living,[5] the Lawton Instrumental Activities of Daily Living Scale,[6] and the Rosow and Breslau scales.[7] Dementia subtypes included AD, vascular dementia, and mixed dementia. Dementia subtyping was based, for AD, on National Institute of Neurological and Communicative Disorders and Stroke–Alzheimer's Disease and Related Disorders Association (NINCDS–ADRDA) criteria, and, for vascular dementia, on National Institute of Neurological Disorders and Stroke–Association Internationale pour la Recherche et l'Enseignement en Neurosciences (NINDS–AIREN) criteria.[8,9] Mixed dementia was defined as diagnosis of AD with either cerebrovascular lesions on brain imaging or a documented history of stroke and presence of prominent executive function deficits in addition to an AD-type cognitive profile.

**Framingham heart study (FHS):** The Framingham Heart Study is a community-based, longitudinal cohort study that was initiated in 1948. The original cohort comprised 5209 residents of Framingham, Massachusetts, and these participants have undergone up to 32 examinations, performed every 2 years, that have involved detailed history taking by a physician, a physical examination, and laboratory testing. In 1971, a total of 5214 offspring of the participants in the original cohort and the spouses of these offspring were enrolled in an offspring cohort. The participants in the offspring cohort have completed up to 9 examinations, which have taken place every 4 years.

All participants have provided written informed consent. Study protocols and consent forms were approved by the institutional review board at the Boston University Medical Center. Surveillance methods have been published previously. Cognitive status has been monitored in the original cohort since 1975, when comprehensive neuropsychological testing was performed. At that time, participants with low cognitive scores (the lowest 10%) also underwent neurologic assessment, and then a dementia-free inception cohort was established that included all dementia-free persons in the entire cohort. Since 1981, participants in this cohort have been assessed at each examination with the use of the Mini–Mental State Examination (MMSE); participants are flagged for further cognitive screening if they have scores below the prespecified cutoffs, which are adjusted for educational level and prior performance. Participants in the offspring cohort have undergone similar monitoring; they answered a subjective memory question in 1979, have undergone serial MMSEs since 1991, and have taken a 45-minute neuropsychological test every 5 or 6 years since 1999. Participants who are identified as having possible cognitive impairment on the basis of these screening assessments are invited to undergo additional, annual neurologic and neuropsychological examinations. If two consecutive annual evaluations show reversion toward normal cognition, participants are returned to the regular tracking pool. Additional examinations are also performed when subjective cognitive decline is reported by the participant or a family member, either spontaneously between examinations or during annual health-status updates; on referral by a treating physician or by ancillary investigators of the Framingham Heart Study; or after review of outside medical records.

A dementia review panel, which includes a neurologist and a neuropsychologist, has reviewed every case of possible cognitive decline and dementia ever documented in the Framingham Heart Study. For cases that were detected before 2001, a repeat review was completed after 2001 so that up-to-date diagnostic criteria could be applied. The panel determines whether a person had dementia, as well as the dementia subtype and the date of onset, using data from previously performed serial neurologic and neuropsychological assessments, telephone interviews with caregivers, medical records, neuroimaging studies, and, when applicable and available, autopsies. After a participant dies, the panel reviews medical and nursing records up to the date of death to assess whether the participant might have had cognitive decline since his or her last examination.[10,11]

The diagnosis of dementia is based on criteria from the Diagnostic and Statistical Manual of Mental Disorders, fourth edition (DSM-IV). The diagnosis of Alzheimer's disease is based on criteria for possible, probable, or definite Alzheimer's disease from the National Institute of Neurological and Communicative Disorders and Stroke and the Alzheimer's Disease and Related Disorders Association (NINCDS–ADRDA). The diagnosis of vascular dementia is based on criteria for possible or probable vascular dementia from the National Institute of Neurological Disorders and Stroke and the Association Internationale pour la Recherche et l'Enseignement en Neurosciences (NINDS–AIREN).[12] The diagnostic algorithm allows participants to have more than one subtype of dementia.

**Cardiovascular Health Study (CHS):** Follow-up for incident dementia was completed as part of the CHS Cognition Study. The study required participants to have undertaken a brain MRI and modified mini-mental state examination (3MSE) between 1992 and 1994. There were 3,660 participants who completed brain MRI and, of these, 3608 also completed the 3MSE and were included in the dementia follow-up cohort. A standardized protocol for dementia surveillance was developed across the four sites, the details of which have been described previously.[13] In brief, the Pittsburgh site endeavored to perform comprehensive neuropsychological testing on all surviving participants. At the three remaining sites, participants at high risk of dementia and minority participants were approached for comprehensive neuropsychological testing and study review. High risk of dementia was defined as one of the following (i) a previous score of less than 80 on the 3MSE, (ii) a decrease of 5 or more points on the 3MSE from any previous examination, (iii) a previous Telephone Interview for Cognitive Status score less than 28, (iv) an Informant Questionnaire on Cognitive Decline in the Elderly score greater than 3.6, (v) incident stroke, or medical record review with dementia diagnosis, or (vi) were residing in a nursing home. Participants with suspected cognitive impairment as identified from follow-up neuropsychological testing underwent a neurological examination. For persons at high risk of dementia who declined further neuropsychological testing or who were deceased, we performed a medical record review of all hospitalizations, questionnaires sent to his/her physician, and standardized interviews by phone with living participants or a designated informant. A committee, comprising a neurologist and psychiatrist from each study site, reviewed all available information to determine a consensus dementia diagnosis. The diagnosis of dementia was based on a progressive or static cognitive deficit of sufficient severity to affect the subjects' activities of daily living (ADL), and a history of normal intellectual function before the onset of cognitive abnormalities. Patients were required to have impairments in two cognitive domains, which did not necessarily include memory.

The neurologists first classified the cases as dementia, MCI, or normal and then, among the dementia cases, by types of dementia. The decision as to whether dementia was present or not was not affected by the results of the MRI. The classification of the specific types of dementia by different categories of classification (e.g. DSM-IV, ADDTC) was done after review of the MRI. Classifications were based on Diagnostic and Statistical Manual of Mental Disorders – ed. 4 (DSM-IV) or National Institute of Neurological and Communicative Diseases and Stroke/Alzheimer's Disease Related Disorders Association (NINDS-ADRDA), State of California Alzheimer's Disease Diagnostic and Treatment Centers (ADDTC) and National Institute of Neurological Diseases and Stroke-Association Internationale pour la Recherche et l'Enseignement en Neurosciences (NINDS-AIREN). [8,14,15]

**Ages Gene/Environment Susceptibility (AGES) - Reykjavik Study:** The AGES-Reykjavik Study is a single center prospective cohort study based on the Reykjavik Study. The Reykjavik Study was initiated in 1967 by the Icelandic Heart Association to study cardiovascular disease and risk factors. The cohort included men and women born between 1907 and 1935 who lived in Reykjavik at the 1967 baseline examination. Reexamination of surviving members of the cohort was initiated in 2002 as part of the AGES-Reykjavik Study. AGES is designed to investigate aging using a multifaceted comprehensive approach that includes detailed measures of brain function and structure. All cohort members were European Caucasians. The study design has been described previously. Briefly, as part of a comprehensive examination, all participants answered a questionnaire, underwent a clinical examination, multiple digital measurements were acquired, and blood was drawn. The dementia case finding was based on a 3-step procedure. All participants were screened on the

Mini-Mental State Examination and DSST. Screen positives on either of the tests were administered another more complete diagnostic test battery. Those screening positive on the Trails A and B or the Rey Auditory Verbal Learning Test went for a final assessment that included a proxy interview and a neurologic examination. The diagnosis of dementia and subtypes was made during a consensus conference that included a geriatrician, a neurologist, a neuropsychologist, and a neuroradiologist who provided a clinical reading of MRI. Dementia was diagnosed according to the guidelines of the DSM-IV. Alzheimer's disease (AD) was diagnosed according to the criteria of the National Institute of Neurological and Communicative Diseases and Stroke–Alzheimer's Disease and Related Disorders Association. Vascular dementia (VaD) was diagnosed following the criteria of the State of California AD Diagnostic and Treatment Centers; Clinical medical history and MRI were used in the diagnosis. It was possible to diagnose a subject with possible AD and possible VaD if the 2 pathologies were thought to contribute to dementia.[16,17]

**EPOZ:** The Epidemiological Prevention Study of Zoetermeer (EPOZ) started in 1975, with the aim of assessing the prevalence of several chronic diseases and their determinants in the city of Zoetermeer.[18] In 1995 and 1996, a random subsample of the participants who were between ages 60 and 90 underwent cognitive testing and brain MRI (N=514); data from this group are considered the baseline for the present study.[19] Participants were screened at study entry and at follow-up visits for dementia, using a strict protocol.

**MEMENTO:** Memento is a French multicenter cohort that aims to improve current knowledge of the natural history of Alzheimer's disease and related disorders (ADRD) and identify new patient phenotypes associated with the risk of developing dementia. The Memento cohort includes patients from the 26 participating memory clinics across France between 2011 and 2014. Participants were followed at least annually for a median of 5 years.[20] Individuals were eligible for inclusion if they (1) were 18 years or older; presented with at least one cognitive deficit defined as performing worse than 1 SD to the mean in one or more cognitive domains (considered as MCI), or (2) presented with an isolated cognitive complaint and were 60 years of age or older. They also had to score on the clinical dementia rating (CDR) scale ≤0.5 (i.e., not demented); have sufficient visual and auditory abilities to partake in neuropsychological testing; and have health insurance, as required by the French government (France has universal access to health care for all legal residents, independent of age, professional standing, or revenue). All participants signed an informed consent form. All incident cases of dementia were reviewed by a panel of expert neurologists/geriatricians, blinded to genetic and biological biomarkers using the Diagnostic and Statistical Manual of Mental Disorders (DSM-IV criteria).[21]

The etiologic diagnosis of dementia was made according to NINCDS-ADRDA for Alzheimer Disease, DLB consortium for dementia with Lewy bodies, Rascovsky criteria for frontotemporal lobar degeneration and NINDS-AIREN for Vascular dementia.[22]

**Rotterdam study (RS):** The Rotterdam Study is a population-based cohort study among inhabitants of a district of Rotterdam (Ommoord), the Netherlands, that aims to examine the determinants of disease and health in the elderly with a focus on neurogeriatric, cardiovascular, bone, and eye disease. In 1990-1993, 7,983 persons aged ≥ 55 years participated and were re-examined every 3 to 4 years (RS1). In 2000-2001, the cohort was expanded by 3,011 persons who were of the same age but had not yet been part of the Rotterdam Study (RS2) and recently moved into the area. In 2006-2008 a second expansion (RS3) of 3,932 persons aged 45 and over was realized. All participants had DNA extracted at their first visit.

Participants were screened for dementia at baseline and subsequent centre visits with the Mini-Mental State Examination and the Geriatric Mental Schedule organic level.[23] Those with a Mini-Mental State Examination score <26 or Geriatric Mental Schedule score >0 underwent further investigation and informant interview, including the Cambridge Examination for Mental Disorders of the Elderly. At each centre visit, all participants also underwent routine cognitive assessment, including a verbal fluency test (animal categories), 15-word learning test, letter-digit substitution task, Stroop test, and Purdue pegboard task. In addition, the entire cohort was continuously under surveillance for dementia through electronic linkage of the study database with medical records from general practitioners and the regional institute for outpatient mental health care. Available information on clinical neuroimaging was used when required for diagnosis of dementia subtype. A consensus panel led by a consultant neurologist established the final diagnosis according to standard criteria for dementia (DSM-III-R) and Alzheimer's disease (NINCDS–ADRDA). Follow-up until 1st January 2016 was virtually complete (96.3% of potential person-years). Within this period, participants were censored at date of dementia diagnosis, death, loss to follow-up, or 1st January 2016, whichever came first.[23]

**UK Biobank:** UK Biobank is a large cohort study of more than 500,000 people recruited from across England, Scotland, and Wales. Strengths of UK Biobank include its size, detailed baseline assessment and measurements including biological samples, follow-up assessments for certain issues, and the availability of long-term linkage to outcome data. The diagnoses of all-cause dementia were obtained using hospital inpatient records from the Hospital Episode Statistics for England, the Scottish Morbidity Record data for Scotland, and the Patient Episode Database for Wales. ICD 9/10 codes mentioned in the table below, were used in the diagnosis of the dementia outcomes. From the baseline (2006-2010) period, the UK biobank participants were followed (2011-2014) up to the earliest incident dementia diagnosis, date of death, the last data collection date by the general practitioner, or the last time of hospital inpatient admission, whichever occurred first.

| ICD9 | | | | ICD10 | | |
|---|---|---|---|---|---|---|
| 331 | 290.20 | *291.2* | *331.5* | G30 | F00.9 | F01.8 |
| 797 | 290.21 | 294.1 | 331.6 | G30.0 | F02 | G31.1 |
| 290 | 290.3 | 294.11 | 331.7 | G30.1 | F02.81 | F01.2 |
| 290.0 | 290.4 | 294.2 | 331.8 | G30.9 | F02.80 | F03 |
| 290.1 | 290.40 | 294.8 | 331.82 | G31.0 | F02.8 | F01 |
| 290.10 | 290.41 | 331.0 | 331.89 | F05.1 | F02.3 | F01.0 |
| 290.11 | 290.42 | 331.1 | 331.9 | F00 | F02.0 | F01.9 |
| 290.12 | 290.43 | 331.11 | 333.0 | F00.0 | F01.3 | |
| 290.13 | 290.8 | 331.19 | *333.4* | F00.1 | F01.1 | |
| 290.2 | 290.9 | 331.2 | | F00.2 | G30.8 | |

**HUNT:** The Trøndelag Health Study (HUNT) is a population-based cohort that has collected samples during four different time periods: HUNT1 (1984-1986), HUNT2 (1995-1997), HUNT3 (2006-2008) and HUNT4 (2017-2019). Participation in HUNT is based on informed consent, and the study has been approved by the Data Inspectorate and the Regional Ethics Committee for Medical Research in Norway (REK: 2014/144). HUNT2 and 3 individuals with baseline information, hospital registry data and genotyping data available were used in the analysis. More detailed description of the genotyped dataset can be found from Brumpton et al.[24,25]

The health care system in Norway is publicly funded. Levanger hospital and Namsos hospital, which are the two only hospitals in Nord-Trøndelag, have catchment area responsibilities for the whole county. Diagnoses of dementia are mainly made at geriatric, neurological, and old age psychiatric wards and outpatient clinics. We obtained data from hospital registries on

ICD-9 and ICD-10 codes from all inpatient and outpatient contacts from 1987 through 2018 for all genotyped participants in the HUNT study. A subset of these diagnoses has been validated against the ICD-10 criteria for Alzheimer's disease by four specialists in geriatrics and old age psychiatry as part of the Health and Memory Study.[26]
Based on ICD-codes (see table under UK Biobank) we defined cases of all-cause dementia, by at least one local hospital contact due to the diagnoses. Incident cases were defined as those who at participation in HUNT had never received any of the diagnoses listed under all-cause dementia, but fulfilled criteria for either of the dementia types during follow-up (till 2018). We defined participants as controls if they had never received any of the diagnoses listed under all-cause dementia during follow-up (until 2018).
**Estonian Biobank (EBB):** The Estonian Biobank (EstBB) is a population-based cohort of 200,000 participants with a rich variety of phenotypic and health-related information collected for each individual.[27] At recruitment, all participants signed a consent to allow follow-up linkage of their electronic health records (EHR), thereby providing a longitudinal collection of phenotypic information. Health records have been extracted from the national Health Insurance Fund Treatment Bills (from 2004), Tartu University Hospital (from 2008), and North Estonia Medical Center (from 2005). The diagnoses are coded in ICD-10 format. For the current study, dementia cases by searching the EHRs for data on the following ICD-10 diagnosis (G30, G30.0, G30.1, G30.9, G31.0, F05.1, F00, F00.0, F00.1, F00.2, F00.9, F02, F02.81, F02.80, F02.8, F02.3, F02.0, F01.3, F01.1, G30.8, F01.8, G31.1, F01.2, F03, F01, F01.0, F01.9). All remaining participants who did not have any of the mentioned ICD-10 codes were defined as controls.
**The Charles F. and Joanne Knight Alzheimer Disease Research Center (Knight ADRC):** The Knight ADRC at Washington University School of Medicine (St. Louis, MO, USA) recruits and longitudinally assesses community-dwelling adults older than 45 years old via prospective studies of memory and aging since 1979. All studies were approved by the Human Research Protection Office at Washington University, and written informed consent was obtained from all participants. The Memory and Aging Project at the Knight ADRC (Knight ADRC-MAP) involves longitudinal collection of biofluids (plasma, CSF, fibroblast), annual clinical assessments, neuropsychological testing, and neuroimaging studies, as well as collection of autopsied brain samples. Eligible participants may be asymptomatic or have mild dementia at the time of enrollment. All participants are required to participate in core study procedures, including annual longitudinal clinical assessments, neuropsychological testing, neuroimaging, and biofluid biomarker studies. Annual assessments of the participants were performed by experienced clinicians using a semi-structured interview with knowledgeable collateral source and the symptomatic individual in accordance with the Uniform Data Set protocol of the National Alzheimer's Coordinating Center, as well as a detailed neurological examination. Participants comprise Non-Hispanic White individuals from North America (82.5%) and African-Americans (13.3%). Samples have been obtained from over 5,510 participants, including 2,426 AD cases, 148 FTD, 88 DLB, and 2,156 cognitively normal healthy individuals. Autopsy material are available for over 1,182 participants including 474 with fresh frozen parietal brain tissue (https://dss.niagads.org/datasets/ng00127/ ). Multi-tissue (brain, CSF, and plasma), multi-omics data (genetics, epigenomics, transcriptomics, proteomics, and metabolomics) have been generated for the purpose of identifying novel risk and protective variants for dementia, and potential drug targets. Participants from the Knight ADRC were included in this study if they were cognitively unimpaired with a global clinical dementia rating (CDR) score of 0 at enrollment. A clinical diagnosis of incident dementia is considered by study clinicians at the conclusion of each annual assessment, integrating results from the clinical assessment and bedside measures of cognitive function.[28] Dementia was diagnosed according to the National Institute of Neurological Disorders and Stroke criteria[8]

and National Institute on Aging-Alzheimer's Association Work Group criteria for participants assessed after 2011.[8] Diagnosis of AD dementia was made in accordance with criteria developed by working groups from the National Institute of Aging and the Alzheimer's Association. [8] Diagnosis of vascular dementia conformed to the NINDS-AIREN criteria.[9]

**Genetic instruments selection**

Genetic variants (SNPs) with minor allele frequency (MAF $\geq 0.01$) that are robustly (genome-wide significant; $P < 5\text{x}10^{-8}$) associated with each of the risk factors [WMH (25 SNPs), stroke (32 SNPs), systolic blood pressure – SBP (472 SNPs), diastolic blood pressure – DBP (477 SNPs), and pulse pressure – PP (449 SNPs)] from the latest and largest GWAS[29-31] satisfying the no measurement error (NOME) assumption[32,33] were considered as instruments for the univariable 2SMR and the individual level genetic risk score (GRS) analysis. Association statistics (effect estimates, standard errors and effect allele) from the European only analysis were extracted for each of the risk factor SNPs and the independence between the SNPs was checked using the 1000 genome European panel as the reference[34] [linkage disequilibrium (LD) r2 < 0.10]. Genetic instruments for the MVMR analysis were constructed by compiling the instruments selected for the univariable analysis and including the corresponding effect estimates from all the exposures included. Further pruning based on their LD dependence (r2 > 0.01) was performed.[35] The strength of the genetic instruments commonly referred to as the F-statistic (Cragg-Donald F statistic) was calculated from the variance explained by the individual SNPs (eTable 1),[36] a value greater than 10 is generally considered as a strong instrument. We additionally report the conditional F-statistic ($F_{TS}$) implemented in the MVMR R package (available through GitHub)[35] which tests the strength of genetic instruments in predicting all the different exposures included in the model (eTable 2).

**Mendelian randomization analyses**

***Primary step:***

Putative causal effect ($\beta_{IVW}$) of the different risk factors on the AD outcomes was estimated, using the inverse-variance weighting (IVW) method, by the weighted sum of the ratios of beta-coefficients from the SNP-outcome associations for each variant (j) over corresponding beta-coefficients from the SNP-exposure associations ($\beta j$). The ratio estimates from each genetic variant were averaged after weighting on the inverse variance (Wj) of $\beta j$ across L uncorrelated SNPs, implemented as an R package RadialMR (available through CRAN repositories).[37]

$$\beta_{IVW} = \frac{\sum_{j=1}^{L} Wj\beta_j}{\sum_{j=1}^{L} Wj} \quad (1)$$

First, the Rücker model-selection framework[37] was applied sequentially. Briefly, the simplest fixed-effect IVW estimates were estimated along with the test (Cochran's Q statistic) for heterogeneity due to genetic instruments exerting an effect on the outcome and exposure through independent pathways (horizontal/directional pleiotropy).

Then, based on the relaxed assumption of presence of SNP pleiotropic effect independent of their strength as instruments (InSIDE),[38] radial MR-Egger regression– a weighted linear regression similar to IVW but without constraining the intercept term– was carried out. In this context, the Egger intercept can be used as an indicator for the presence of average pleiotropic effects ($P < 0.05$). Next, we tested the relative goodness of fit of the MR-Egger over the IVW approach using a heterogeneity ratio statistic ($Q_R$) that was computed by dividing the MR-Egger heterogeneity (Rücker's Q') statistics over the standard Cochran's Q. $Q_R$ values close to

1 indicates that MR-Egger is no better fit than the IVW. Under the presence of significant heterogeneity (Q-PHet < 0.05), IVW estimates from the random effects model were used and the IVW was repeated after filtering the pleiotropic outliers.

***Pleiotropy – accounting step (Uncorrelated pleiotropy):***

Second, other methods accounting for uncorrelated pleiotropy (horizontal pleiotropic effects directly on the outcome that are uncorrelated with effects on the exposures) such as the weighted median and weighted mode estimators[39] were carried out. Weighted median estimator uses the weighted median and the weighted mode uses the most frequent of the ratio estimates, as opposed to the mean used by the IVW approach. The results from all four of these MR methods (IVW, MR-Egger, weighted median and weighted mode) were compared for consistency thereby strengthening the causal evidence.[40] Third, with the InSIDE assumption satisfied, a robust adjusted profile score method called the RAPS was applied[41] This method, by making the weight that each variant receives a function of the causal effect and instrument strength, provides causal estimates more robust to weak instruments and allows the inclusion of genetic instruments well below the conventional GW significant threshold.[38] A p-value < 0.017 correcting for 3 independent traits was considered significant for the univariable MR.

***Pleiotropy – accounting step (Correlated pleiotropy):***

Additionally, correlated pleiotropic (pleiotropic effects on the outcome that are correlated with effects on the exposures through unmeasured confounders) effects were examined using CAUSE[42] in order to differentiate causal effects from correlated pleiotropy due to unknown or unmeasured shared factors. Briefly, in causal models, all genetic variants with nonzero effect exhibit correlation, whereas in models with shared effects correlation is induced only for a subset of variants. CAUSE models the proportion of variants with non-zero effect from the genome-wide summary statistics; it tests two nested models (model-1: null vs sharing, model-2: null vs causal) and compares them (model-3: sharing vs causal) using a Bayesian model comparison approach while providing an additional indication on the proportion of variants (CAUSE-q) acting through shared factor. CAUSE improves the power of MR analysis by including a larger number of LD-pruned SNPs (LD r2 < 0.10) with an arbitrary $p \leq 1.00E\text{-}03$. CAUSE is a data-driven approach that assumes the true causal model is contributed by all the genetic instruments while the shared-pleiotropic model is influenced only by a subset of the instruments. CAUSE compares the prediction accuracy (expected log pointwise posterior density, ELPD) of the Bayesian model for the causal effect ($ELPD_C$) and the shared effect ($ELPD_S$). A difference ($\Delta ELPD = ELPD_C - ELPD_S$) greater than zero indicates that the posteriors from the causal model predict the data better than the sharing model. Following the analytic scheme (eFigure-1), for the exposures with probability for a better fit of the causal model ($\Delta ELPD > 0$) but with significant evidence of effects due to shared genetic factors (q p-value < 0.05), we additionally validated the causal association using multivariable Mendelian randomization (MVMR) conditioning for effects from specific related traits.

***Multivariable Mendelian randomization:***

MVMR provides direct effect estimates of the exposure of interest by conditioning on every other exposures included in the model. Unlike a conventional MR, careful application of MVMR could ameliorate any selection bias that is common when studying aging related disorder as a result of surviving on the genetically predicted exposure and competing risk of the outcome. The putative relationship between a given exposure and the outcome was explored adjusting for major causes of survival through established risk factors (BP, Stroke, CAD) and/or genetic horizontal pleiotropy. Different combinations of the exposures was considered and the $F_{TS}$ conditional on the other exposure was calculated as a measure of instrument strength (eTable 2).[35] Briefly, MVMR, by regressing a given exposure instrumental variable on all the remaining exposures as controls, generates a predicted value for that exposure that is not correlated with potential confounders and pleiotropic effects. The direct effect estimates from MVMR by adjusting for other main determinants of survival and outcome can be used to address potential collider bias due to selection, as a result of conditioning on the selection on survival of both the genetic instruments (eg. high BP, stroke risk) and common causes of the outcome. Genetic instruments for coronary artery disease (CAD)[43] was additionally used as an exposure in the MVMR analyses. Excessive heterogeneity (PHet < 0.05) in the MVMR causal effect estimates as a result of pleiotropy was measured using the modified version of Cochran Q statistics giving an upper bound on the bias suggesting certain genetic instruments being invalid as a result of weak $F_{TS}$ and possible residual pleiotropy.[44] In MR approaches, weak instrument bias typically leads the causal effect in the direction towards the null but does not overestimate it. Using the Qhet-MVMR function (https://rdrr.io/github/WSpiller/MVMR/man/qhet_mvmr.html), we additionally confirmed on the effect direction (betaQivw) for exposures with significant (P < 0.013, for 4 independent traits) MVMR association after accounting for potential inclusion of weak instruments ($F_{TS}$ < 10) that is estimated by conditioning the effects of the primary exposure with a secondary exposure. We additionally conducted two-sample bidirectional MR within the exposures included in the MVMR for determining the causal relation between the exposures, where genetic instruments for the exposure are tested for a putative causal association with a given outcome and vice versa.

**Longitudinal analyses**

Individual level association of risk scores with all dementia and mortality were performed in the CHARGE cohorts (Three-city study: 3C, Framingham heart study: FHS, Rotterdam study: RS, Age, Gene/Environment Susceptibility - Reykjavik study: AGES, Cardiovascular health study: CHS, Epidemiological Prevention Study of Zoetermeer: EPOZ) the Knight ADRC cohort and different biobanks (HUNT, Estonian biobank and UK biobank).

Genetic instruments constructed for the 2SMR analysis were used to compute weighted genetic risk scores (wGRS) for the different exposures (WMH, stroke, BP traits) in each CHARGE cohort, which were rescaled so that each unit increase in wGRS[45] would correspond to one additional risk allele via the following formula:

$$\mathrm{wGRS_i} = \frac{N}{\sum_{j=1}^{N} \beta_j} \sum_{j=1}^{N} \beta_j d_{ij} \qquad (2)$$

with $\beta_j$ the effect size (or log odds ratio) of the risk increasing allele of the j-th SNP ($SNP_j$) estimated, *N* the number of SNPs selected as the genetic instrument for a given exposure; $d_{ij}$ its dose (genotyped or imputed) for the subject i. All the selected genetic instruments were available and used in calculating wGRS. The wGRSs were standardized to have mean 0 and variance 1.

Analyses were restricted to participants of European ancestry with at least one follow-up visit and no dementia at baseline. The relation between the genetic risk score for a given exposure and incident dementia was explored using Cox models with birth as time origin and age as the time scale in order to avoid the non-proportionality in dementia risk with age. Baseline age was the age at which participants entered the cohort. Follow-up time ended at age of dementia diagnosis for cases or age at the last confirmed dementia-free visit for controls. Analyses were controlled for gender, principal components for population stratification, study-specific criteria, and for education level (eTable 3 in Supplemental 2). Individual cohort-specific estimates were combined using a fixed-effects inverse variance weighted meta-analysis, implemented as an R (meta) package. We presented Hazard ratios (HR) and their 95% confidence interval (CI).

We performed a set of secondary analysis in two large population-based studies (3C Ncases: 621, Ncontrols:4954 and AGES Ncases:978, Ncontrols: 2937), with similar age range and identical dementia ascertainment procedure. To explore whether a possible survival bias during follow-up might affect our results, we built illness death models (IDMs), a type of multistate models, that describe the pathway from an initial state (e.g., alive and without disease) to an absorbing state representing a terminal event (e.g., death) either directly or through an intermediate state (e.g., disease). IDMs can account for interval-censoring of time-to-onset of dementia (related to the fact that diagnosis of dementia is made at intermittent follow-up visits only), and for competing risk of death by modeling the probability of developing the disease between the last visit and death. Herein we jointly modelled all cause dementia onset and death, simultaneously estimating to which extent they were affected by the wGRS in dementia-free participants. IDM were controlled for gender, study-specific criteria, and for education level; we did not adjust for principal components for population stratification due to computational challenges related to IDM (moreover, those variables were not associated with dementia). IDM estimates were than compared with the standard Cox models.[46]

Additionally, to test the effect of polygenicity involved in the primary association results at the individual level, we extended the risk score approach by including genetic instruments for the risk factors from different association levels (p-values <0.5, <0.10, <1E-03,<1E-04,<1E-06, 5E-08) in the original GWAS. Risk alleles were coded as alleles associated with increasing trait values/risk, and only variants with MAF $\geq$ 0.01 were considered. SNPs were first pruned for LD independence at an $r^2$ threshold of 0.2 and a maximum distance of 250 kilobases. A total of 30 polygenic profile score (PGS) (5 risk factor phenotypes at 6 p-value thresholds) were created, assuming additive contributions and simple linear scoring (implemented in PLINK v1.90b3.46). Their associations with incident dementia outcomes (including AD and vascular or mixed dementia) were examined using Cox-models (with birth as time origin and age as the time scale) controlled initially for gender, study-specific criteria, principal components for population stratification, and education level. Further adjustment for interim incident stroke (after excluding individuals with prevalent stroke) was also made. When evaluating one particular subtype of dementia, participants developing other type were censored at their age of dementia. A p-value < 0.017 correcting for 3 independent traits was considered significant. All analyses were performed using SAS software version 9.4 (SAS Institute Inc., Cary, NC, USA) and R statistical software (version 3.3.2, R development core team).

**eAppendix 1. Acknowledgments**

FHS: We thank the study participants, as well as the study team (especially the investigators and staff of the neurology team) for their contributions and dedication to the study. The authors are pleased to acknowledge that the computational work reported on in this paper was performed on the Shared Computing Cluster which is administered by Boston University Research Computing Services. URL: www.bu.edu/tech/support/research/.

CHS: This CHS research was supported by NHLBI contracts HHSN268201200036C, HHSN268200800007C, HHSN268201800001C, N01HC55222, N01HC85079, N01HC85080, N01HC85081, N01HC85082, N01HC85083, N01HC85086, 75N92021D00006; and NHLBI grants U01HL080295, R01HL087652, R01HL105756, R01HL103612, R01HL120393, and U01HL130114 with additional contribution from the National Institute of Neurological Disorders and Stroke (NINDS). Additional support was provided through R01AG023629 from the National Institute on Aging (NIA). A full list of principal CHS investigators and institutions can be found at CHS-NHLBI.org. The provision of genotyping data was supported in part by the National Center for Advancing Translational Sciences, CTSI grant UL1TR001881, and the National Institute of Diabetes and Digestive and Kidney Disease Diabetes Research Center (DRC) grant DK063491 to the Southern California Diabetes Endocrinology Research Center.

AGES: We thank the study participants for their participation. The study was funded by the National Institute on Aging (NIA) (N01-AG-12100, and HHSN27120120022C), Hjartavernd (the Icelandic Heart Association), and the Althingi (the Icelandic Parliament), with contributions from the Intramural Research Programs at the NIA The study was approved by the Icelandic National Bioethics Committee (VSN: 00-063) and the MedStarResearch Institute (project 2003-145).

MEMENTO: The MEMENTO cohort is sponsored by Bordeaux University Hospital (coordination: CIC1401-EC, Bordeaux) and was funded through research grants from the Foundation Plan Alzheimer (Alzheimer Plan 2008–2012), the French ministry of research and higher education (Plan Maladies Neurodégénératives (2016–2019)).

HUNT: The Trøndelag Health Study (HUNT) is a collaboration between HUNT Research Centre (Faculty of Medicine and Health Sciences, NTNU, Norwegian University of Science and Technology), Trøndelag County Council, Central Norway Regional Health Authority, and the Norwegian Institute of Public Health. The genotyping in HUNT was financed by the National Institutes of Health; University of Michigan; the Research Council of Norway; the Liaison Committee for Education, Research and Innovation in Central Norway; and the Joint Research Committee between St Olavs hospital and the Faculty of Medicine and Health Sciences, NTNU. The genetic investigations of the HUNT Study is a collaboration between researchers from the K.G. Jebsen Center for Genetic Epidemiology, NTNU and the University of Michigan Medical School and the University of Michigan School of Public Health. The K.G. Jebsen Center for Genetic Epidemiology is financed by Stiftelsen Kristian Gerhard Jebsen; Faculty of Medicine and Health Sciences, NTNU, Norway.

**eAppendix 2. Conflict of Interest Disclosures**: Dr. Bruce Psaty serves on the Steering Committee of the Yale Open Data Access Project funded by Johnson & Johnson.

eFigure 1: Analytical pipeline for the MR approaches. WMH White matter hyperintensity burden, BP Blood pressure, AD clinically diagnosed late-onset Alzheimer's disease, ADmeta (parental dementia status + AD), ΔELPD Expected log pointwise posterior density, q p-value significance of the sharing (q) model.

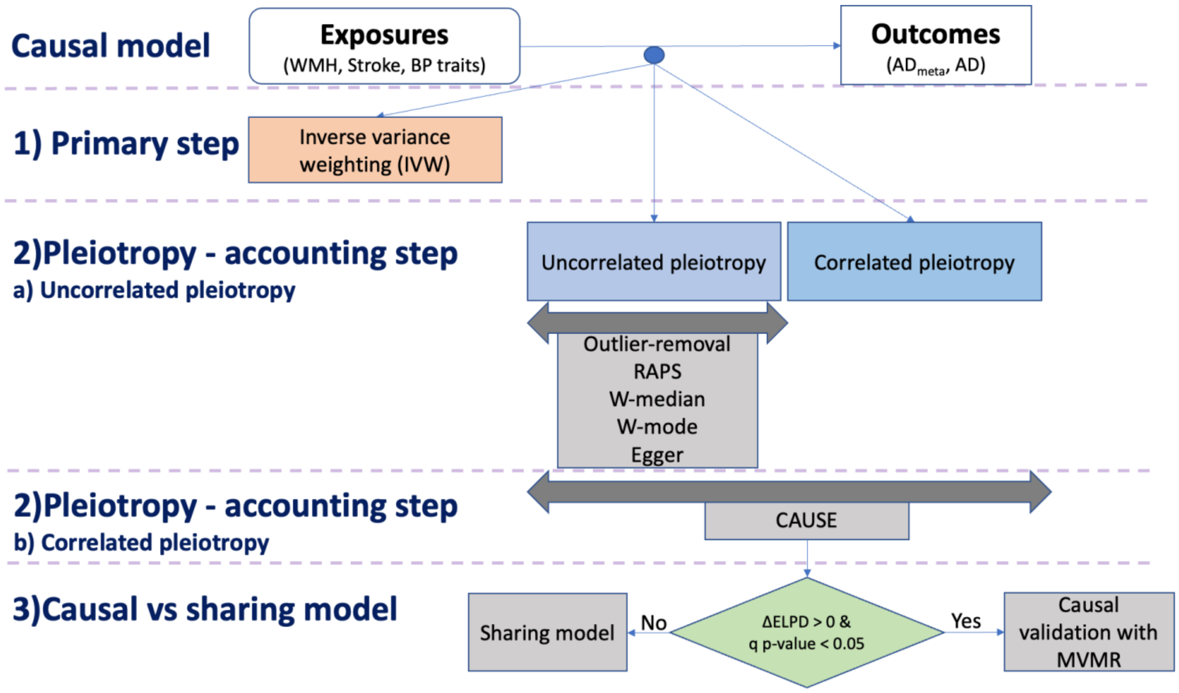


eFigure 2: Mendelian randomization model with different types of pleiotropy; and the included exposures and outcomes. WMH White matter hyperintensity burden. BP Blood pressure, AD clinically diagnosed late-onset Alzheimer's disease, ADmeta (parental dementia status + AD)

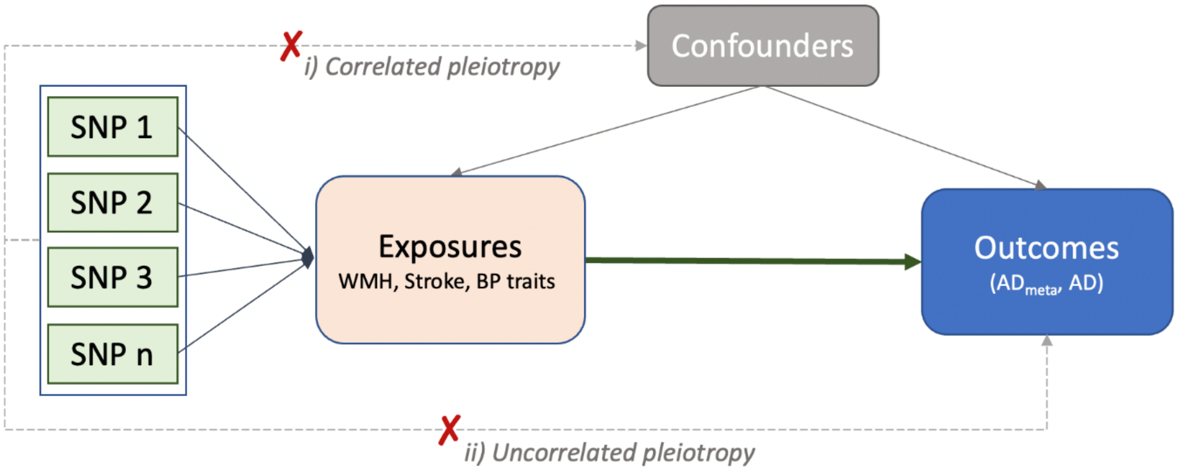

eFigure 3: Mendelian randomization results of vascular risk factors with AD. Point estimates and confidence intervals from the inverse-variance weighted (IVW) method are shown. WMH White matter hyperintensity burden. SBP systolic blood pressure, DBP diastolic blood pressure, PP pulse pressure, RAPS Robust adjusted profile score, W-mode Weighted mode, W-median Weighted median

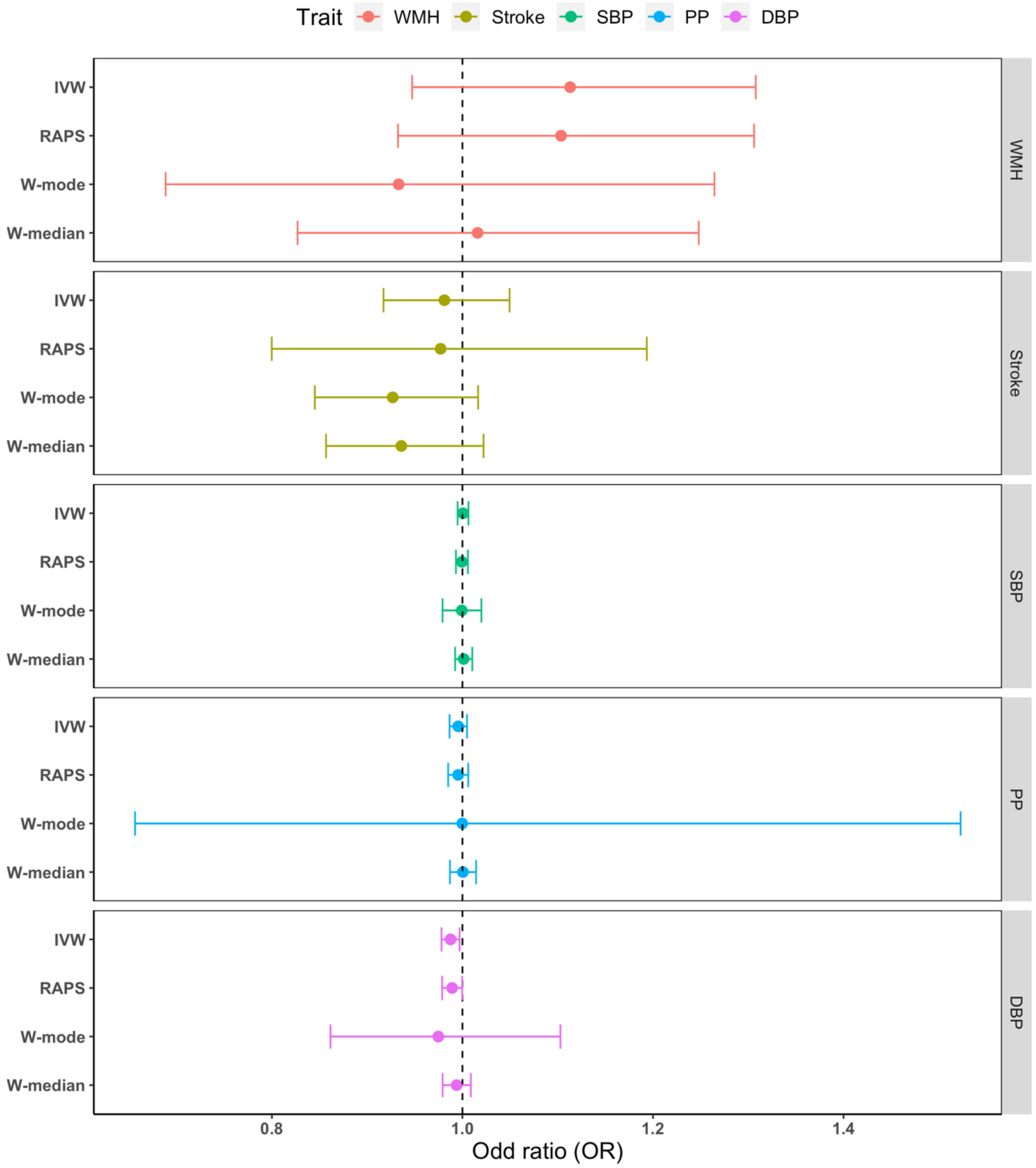

eFigure 4: Comparison of the sharing and the causal model using CAUSE for ADmeta as the outcome. Eta, η: Effect estimate sharing model, η p-value for the shared effect, q: proportion of variants exhibiting correlated pleiotropy, q p-value: p-value for q, Gamma, $\gamma$: Effect estimate causal model, $\gamma$ p-value for the causal effect, ΔELPD: expected log pointwise posterior density, testing causal vs sharing model. ΔELPD > 0 indicates better fit for the causal model.

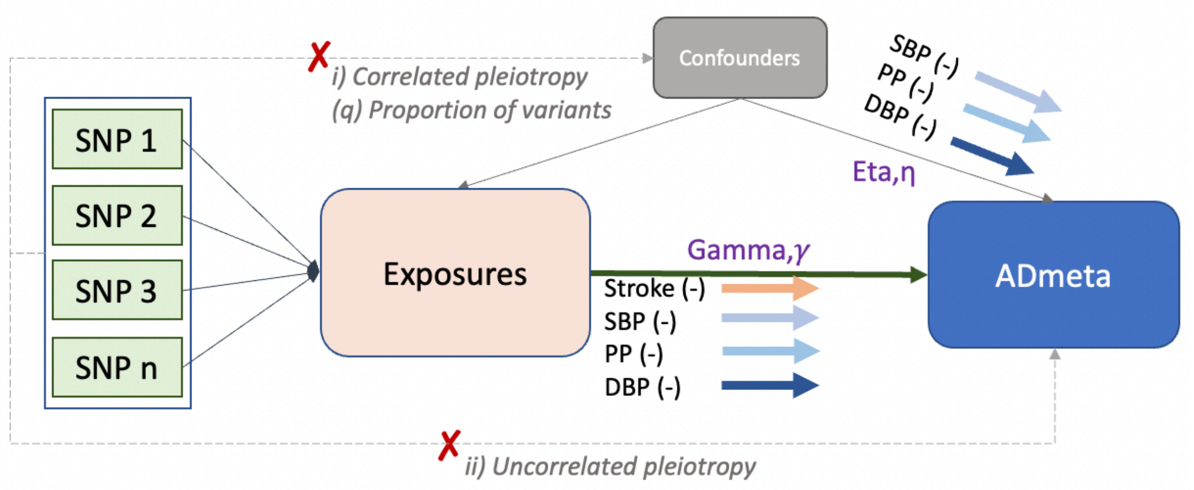


| | Sharing model | | | | Causal model | | Sharing vs Causal |
|---|---|---|---|---|---|---|---|
| Exposure | Eta, η | η p-value | q | q p-value | Gamma,$\gamma$ | $\gamma$ p-value | ΔELPD |
| WMH | 0.19 | 7.99E-01 | 0.03 | 6.22E-01 | 0.04 | 3.91E-01 | 0.50 |
| Stroke | -0.27 | 3.50E-01 | 0.11 | 2.72E-01 | -0.10 | 5.15E-03 | -2.60 |
| SBP | -0.02 | 2.84E-14 | 0.41 | 1.58E-06 | -0.01 | <1.00E-15 | -3.00 |
| PP | -0.04 | 1.01E-04 | 0.29 | 1.21E-03 | -0.01 | 1.01E-04 | -1.60 |
| DBP | -0.04 | 2.74E-07 | 0.39 | 8.85E-06 | -0.02 | 2.84E-14 | -2.20 |

eFigure 5: Comparison of the sharing and the causal model using CAUSE for AD as the outcome. Eta, η: Effect estimate sharing model, η p-value for the shared effect, q: proportion of variants exhibiting correlated pleiotropy, q p-value: p-value for q, Gamma, $\gamma$: Effect estimate causal model, $\gamma$ p-value for the causal effect, ΔELPD: expected log pointwise posterior density, testing causal vs sharing model. ΔELPD > 0 indicates better fit for the causal model.

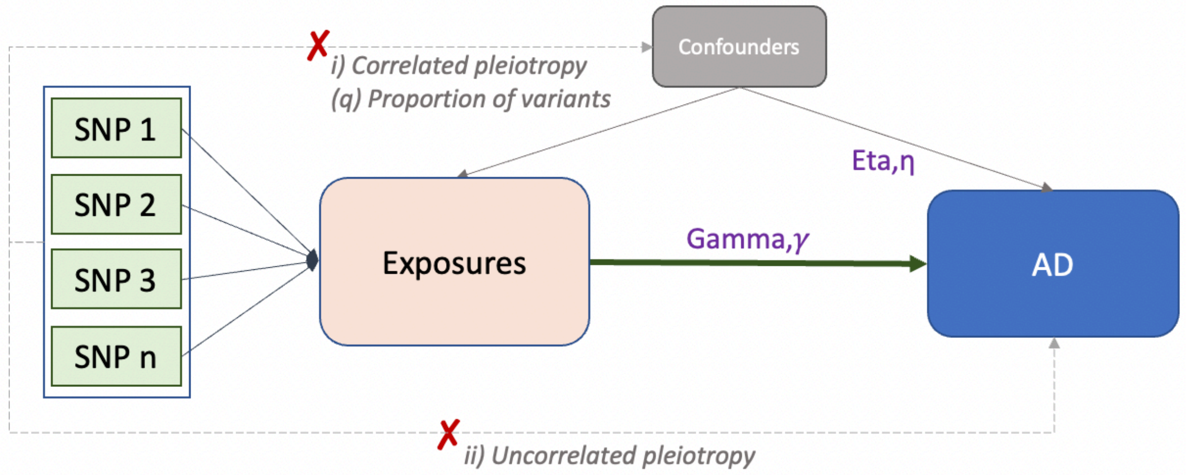


| | Sharing model | | | | Causal model | | Sharing vs Causal |
|---|---|---|---|---|---|---|---|
| Exposure | Eta, η | η p-value | q | q p-value | Gamma,γ | γ p-value | ΔELPD |
| WMH | 0.39 | 7.33E-01 | 0.03 | 8.37E-09 | 0.02 | 7.67E-01 | 0.91 |
| Stroke | -0.21 | 7.34E-01 | 0.04 | 5.24E-01 | -0.04 | 4.17E-01 | 0.41 |
| SBP | -0.04 | 8.11E-02 | 0.09 | 1.59E-01 | 0.00 | 1.00E+00 | 0.44 |
| PP | -0.05 | 4.41E-01 | 0.05 | 4.22E-01 | 0.00 | 1.00E+00 | -0.97 |
| DBP | -0.05 | 7.44E-02 | 0.14 | 8.60E-02 | -0.01 | 4.96E-02 | -1.20 |

eFigure 6: Forest plot showing association of WMH-wGRS with incident all-cause dementia (study-wise). Results are for the main model adjusting for gender, principal components for population stratification, study-specific criteria, and education level. Studies with memory clinic study design: MEMENTO.

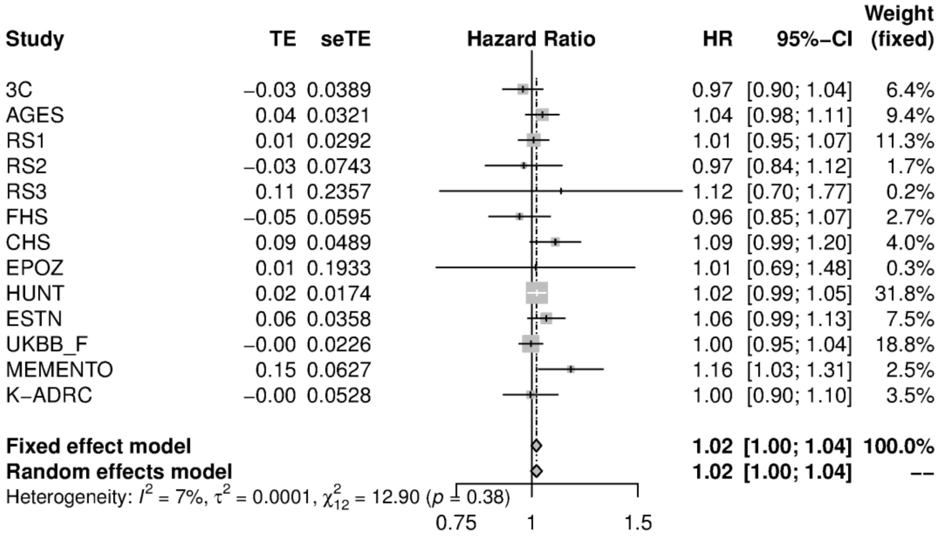

eFigure 7: WMH polygenic profile score (PGS) association with dementia outcomes in 3C (blue) and AGES (black). Left plot: Main analysis, Right plot: Sensitivity analysis after adjusting for interim stroke. P-value selection thresholds: p-value < 5.00E-08 (S1), <1.00E-06 (S2), <1.00E-04 (S3), <1.00E-03 (S4), <0.10 (S5), <0.50 (S6).

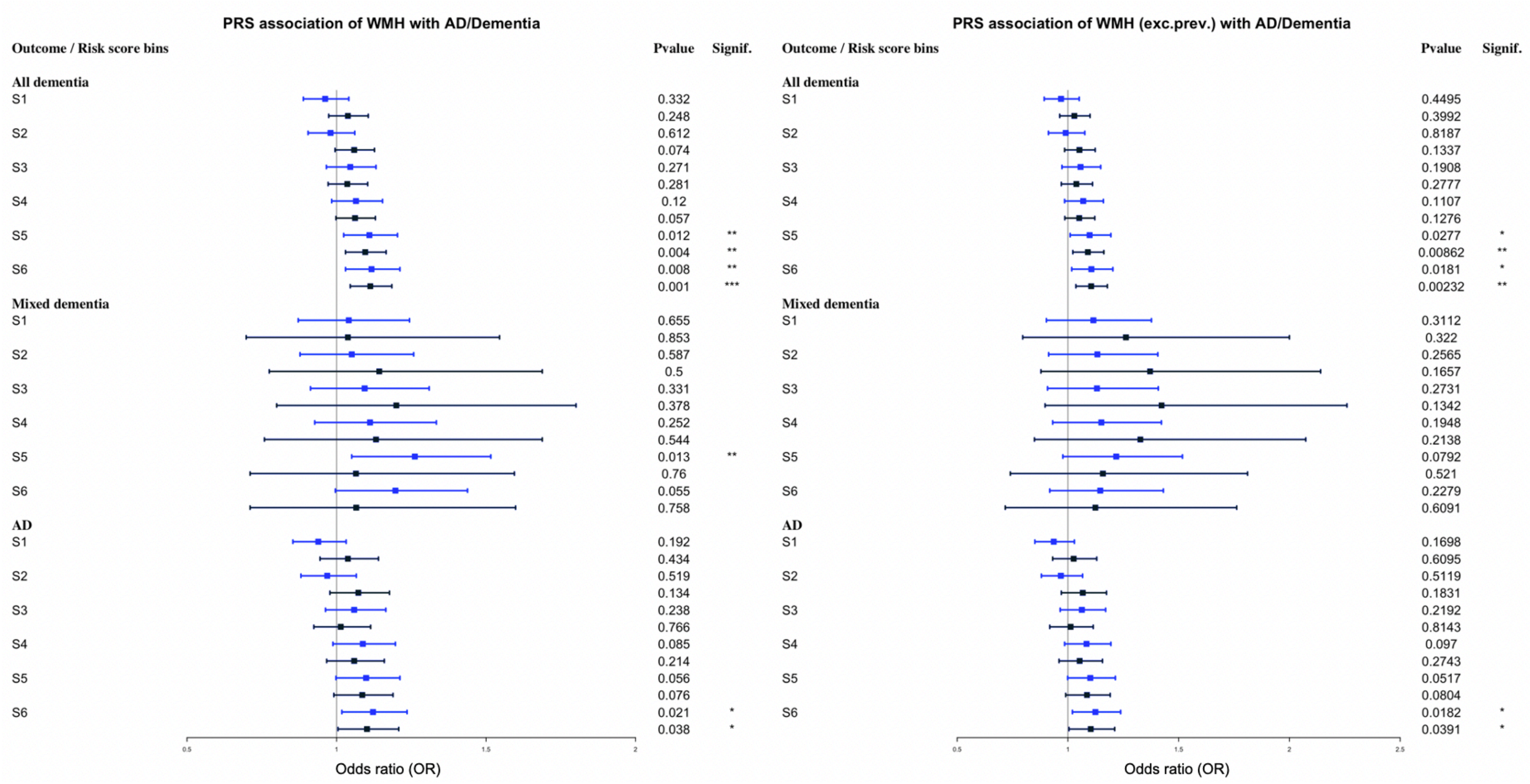

eFigure 8: Stroke polygenic profile score (PGS) association with dementia outcomes in 3C (blue) and AGES (black). Left plot: Main analysis, Right plot: Sensitivity analysis after adjusting for interim stroke. p-value < 5.00E-08 (S1), <1.00E-06 (S2), <1.00E-04 (S3), <1.00E-03 (S4), <0.10 (S5), <0.50 (S6).

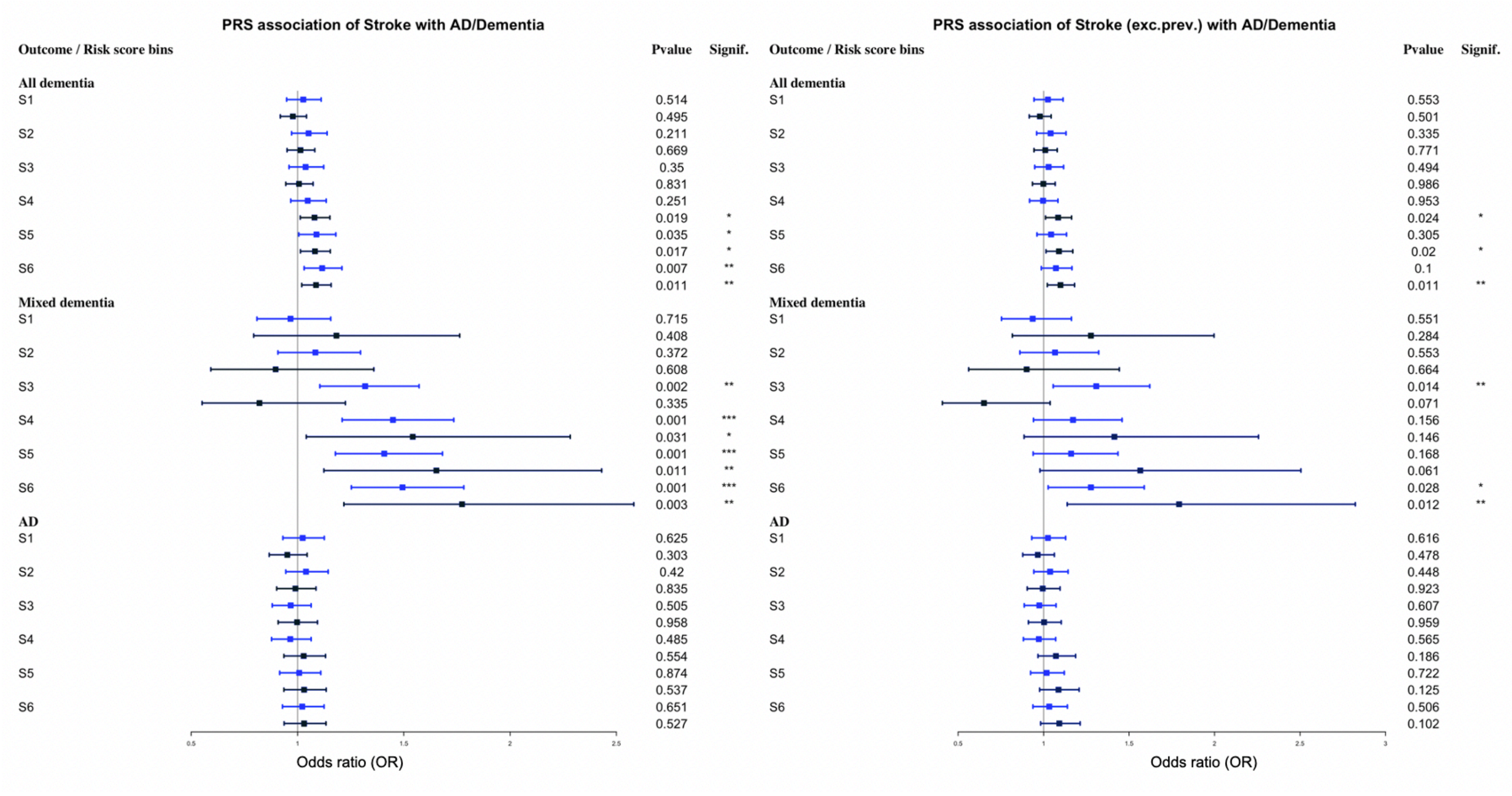
